# Measurement of the extent of Anxiety and Depression that has occurred in college students due to the COVID 19 pandemic: A Survey based cross-secConal study

**DOI:** 10.1101/2021.12.18.21268026

**Authors:** Shubham Goswami, Soujanya Chakraborty, Aritra Chakraborty

**Affiliations:** Bankura Sammilani Medical College and Hospital, Bankura, West Bengal; Department Of Psychiatry, Bankura Sammilani Medical College and Hospital, Bankura, West Bengal

## Abstract

**OVERVIEW:** The ongoing Pandemic because of the Coronavirus disease 2019 (COVID-19) has caused all the educational institutes including colleges to be closed for a very long time. As a result the students are compelled to remain in their homes for this time. Prolonged stay at home along with excess use of social media and other modes to “kill” the time are quite famous to cause certain health issues in a person, specially the teenagers and adolescents. Mental wellbegin, being a dimension of health as per WHO should not be ignored at all specially in these situations.

**METHOD OF STUDY:** An Online Questionnaire is prepared based of the ZUNG Self Rating Anxiety and Self Rating Depression Scale (Pre-validated Scales). The Form is circulated digitally among the people and then we have collected the data in excel. Based on the result we have prepared our statistical chart.

**RESULT:** Quite a significant number of candidates were suffering due to the pandemic situation. 17.091% were suffering from mild to moderate anxiety, 1.785% had marked to severe anxiety levels(Constituting approximately 18.9% of the total). On the other hand, 8.673% of the students had mild depression, while 1 candidate (0.255%) had moderate depression and 1 (0.255%) had severe depression, (Constituting approximately 9.20% of the total). We found that candidates in the age group of 23-24 years had the maximum prevalence of depression, it was followed by candidates with age between 21-22 years. We found that the candidates with age between 23 to 24 years were having highest prevalence of significant anxiety levels which is closely followed by candidates having age which lies between 22 years to 23 years.

## Introduction

^**1**^ The World Health Organization (WHO) defines health as “A state of complete physical, mental and social wellbeing and not merely the absence of disease or infirmity.”

Mental health is still considered a stigma, and millions of them are silently suffering from it without any external help or support.

Adolescents, especially college-going students, suffer from both depression and anxiety at higher rates as the stressors/triggers are present in abundance. ^**2**^ Emotional, behavioral, sexual, economic, academic, and social changes and efforts of discovering one’s identity with psychosocial and maturation also occur. ^**3**^ They go through critical transitory period in their life in which they are going from adolescence to adulthood making major life decisions. During this period, the mental health of university youth constitutes one of the important components of social health. It is known to affect an individual’s academic performance greatly. ^**4**^ According to the WHO report, practically, all psychiatrists who have had extensive experience in working with college students agree that about 10% of the members of any institution of higher learning are likely to have emotional problems at some time or other during each year which interferes seriously with their work. ^**5**^ Depression is under-recognised among adolescents because depressive symptoms are considered a familiar part of adolescent experience. These triggers often exert ^**6**^ a negative effect with catastrophic consequences on students’ academic performance, physical health, and psychological well-being. Provided in the current situation of Pandemic in which students are forced to stay in their homes since a very long time along with delay in their academic curriculums, they are more prone to get affected by mental health issues like anxiety and depression. Hence it is the need of an hour to see the situation of the students due to this pandemic.

## Aims and Objective

### General

The purpose of this study was to measure the extent of anxiety and depression among the students of different colleges throughout the India during COVID-19 pandemic and to investigate the prevalence of depression and anxiety among them. It also aimed at identifying the determinants of depression and anxiety. This would help us in assessing the prevalence and the current status of anxiety and depression among the college students by generating a standardised e-questionnaire using the Google Form, and sharing the link through social media. We can also assess the risk of causing anxiety and depression by throwing light on the knowledge-gap regarding these psychiatric illnesses.

### Specific

1. To measure the extent of anxiety and depression among the college-going students that has occurred during COVID 19 pandemic.
2. To assess the risk or prevalence of anxiety and depression among the students during pandemic.

## Methodology

This is a Online Questionnaire based study which is done in India. The questionnaire is made on the lines of prevaildated [Zung Self-Rating Anxiety Scale](https://www.mnsu.edu/comdis/isad16/papers/therapy16/sugarmanzunganxiety.pdf) and [Zung Self-Rating Depression Scale](http://www.mentalhealthministries.net/resources/flyers/zung_scale/zung_scale.pdf). Along with this a few questions to asses the Socioeconomic status, Age, Residence were also added. The data is taken in excel; sheet and statistical graph is prepared using SPSS.

The questions added in addition to prevalidated scales are:-

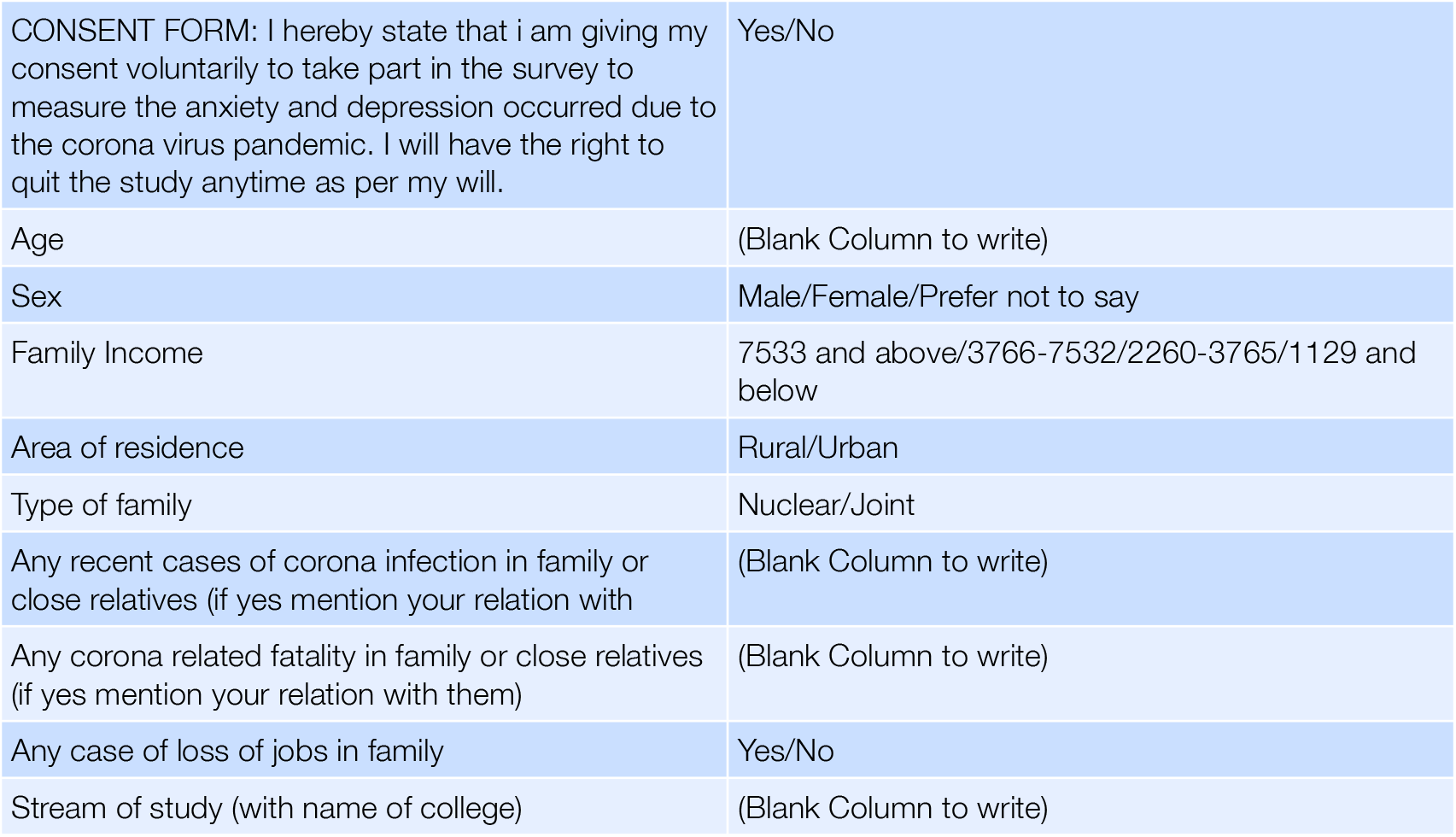

|  |  |
| --- | --- |
| CONSENT FORM: I hereby state that i am giving my consent voluntarily to take part in the survey to measure the anxiety and depression occurred due to the corona virus pandemic. I will have the right to quit the study anytime as per my will. | Yes/No |
| Age | (Blank Column to write) |
| Sex | Male/Female/Prefer not to say |
| Family Income | 7533 and above/3766-7532/2260-3765/1129 and below |
| Area of residence | Rural/Urban |
| Type of family | Nuclear/Joint |
| Any recent cases of corona infection in family or close relatives (if yes mention your relation with | (Blank Column to write) |
| Any corona related fatality in family or close relatives (if yes mention your relation with them) | (Blank Column to write) |
| Any case of loss of jobs in family | Yes/No |
| Stream of study (with name of college) | (Blank Column to write) |

### Data source

The survey was conducted from 15 October 2021 to 15 November 2021. Students enrolled in different colleges across India were the target population. An easy to understand questionnaire was used to collect ‘basic information,’ ‘depression,’ and ‘anxiety’ related information. An online-based platform was used to distribute the e-questionnaire, developed by using the Google Form, to the students. College students from all the divisions in India were contacted through different social networks and interviewed.

#### Study Type

A Cross-sectional type of analytical observational study.

### Timelines

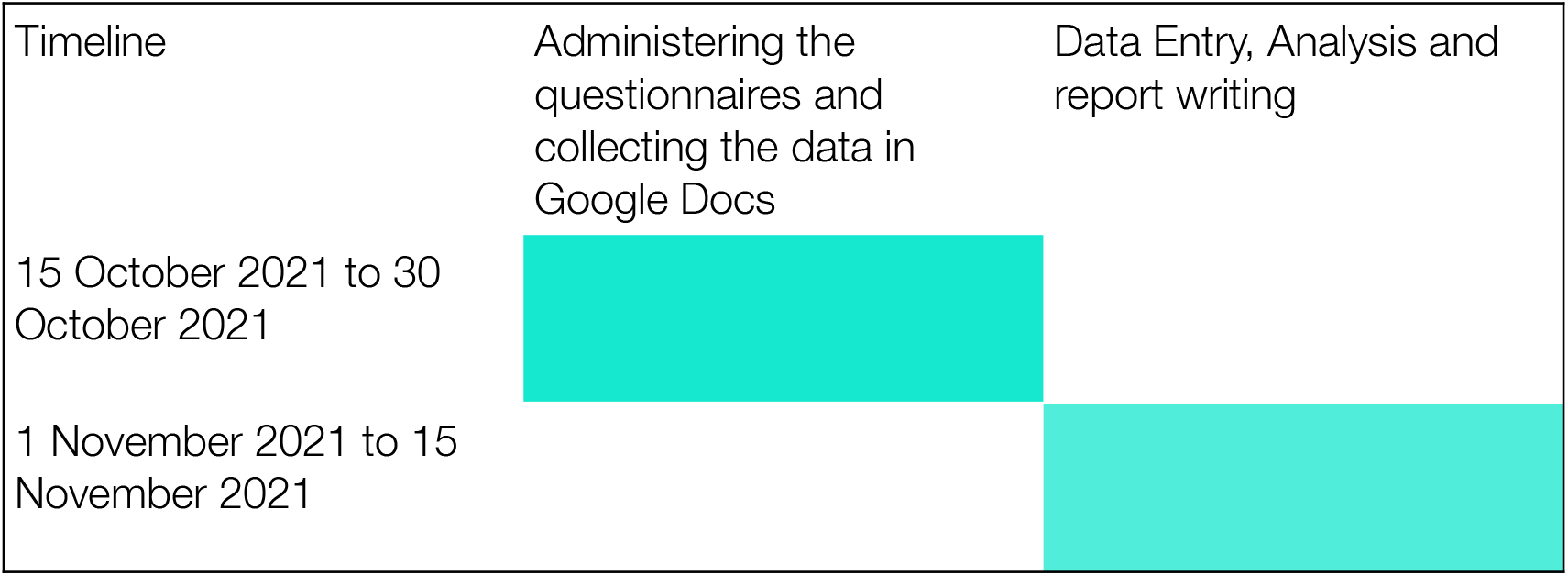

#### Place of Study

This is a virtual study on different social media platforms, among the students of different colleges across the India.

#### Duration of Study

Two months

#### Study Population

All Undergraduate, graduate, and postgraduate college-going students of any age group but exposed to the curriculum for at least 1 year.

### Sample Size and Sampling Technique

#### Sample size

According to AISHE 2020, total number of enrolment in higher education has been estimated to be 38.5 million. Considering it as population size, the sample size was calculated with the help of Raosoft Sample size calculator. Using a margin of error of 5%, confidence level of 95%, and response distribution of 50%, the sample size came out as 385. Considering 10% Non-Response rate (NRR), the size stands 428 (427.7).

#### Sampling Technique

Purposive sampling technique till the targeted sample size is reached.

### Inclusion criteria

Undergraduate, graduate, and postgraduate students of any age group but exposed to the curriculum for at least 1 year were included in the study.

### Exclusion criteria

Students who were unwilling to participate, who submitted incomplete data and those who did not come under the given inclusion criteria were excluded from the study. Microsoft Excel was used for calculating the findings and results.

### Study Tools

1. Administered questionnaire (Pre-validated for the purpose of the study)
2. Google Sheet

### Procedure / Study Technique

To determine the prevalence or to measure the extent of anxiety and depression level among the students, first they were registered or enlisted in a Google sheet. During the course of enlistment an informed consent was taken & only those who gave the consent, were included in the study. A pre-validated Questionnaire on anxiety and depression was be supplied to each student virtually which was filled by them.

Data was collected with the help of the questionnaire consisting of sociodemographic data and the above mentioned scales. The forms was majorly distributed and forwarded digitally through Google Forms.

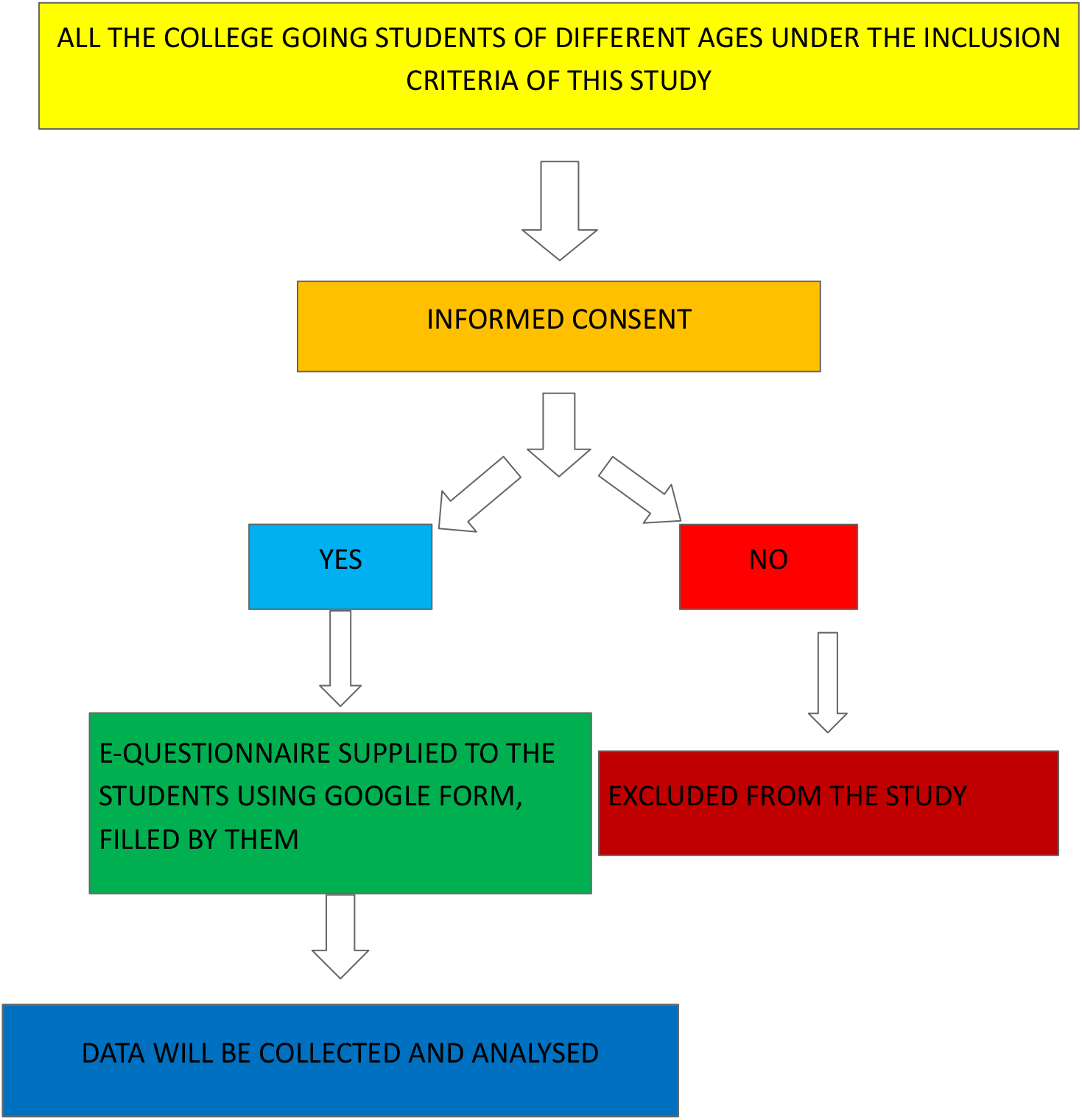

### Study Variables

#### Demographic profile

1. Age
2. 2. Sex
3. Religion,
4. Education status,
5. Residence (urban/ rural),
6. Income (above poverty line/ below poverty line)

#### Anxiety and Depression

1. Physical health
2. Psychological health
3. Social relationship
4. Environment

## DATA COLLECTION AND ETHICAL CLEARANCE

This study was conducted after due getting permission from Institutional Ethics Committee (IEC). All college-going students will be included in the study based on inclusion and exclusion criteria. A total of 392 students were included in the study (Which is a value in between 385 and 428)

## RESULT

We collected data from 392 students from around the various states of India, who belongs from various age groups.

| Age group | Number of students in study |
| --- | --- |
| 18-19 | 11 |
| 19-20 | 53 |
| 20-21 | 128 |
| 21-22 | 105 |
| 22-23 | 59 |
| 23-24 | 18 |
| 24-25 | 13 |
| 25-26 | 2 |
| 26-27 | 2 |
| 29 | 1 |
| TOTAL | 392 |

### 1. SEX

Our study had 56.6% male and 42.9% female participants, this ensures both the gender are nearly equally represented. Rest 0.5% of the participants preferred not to disclose their gender.

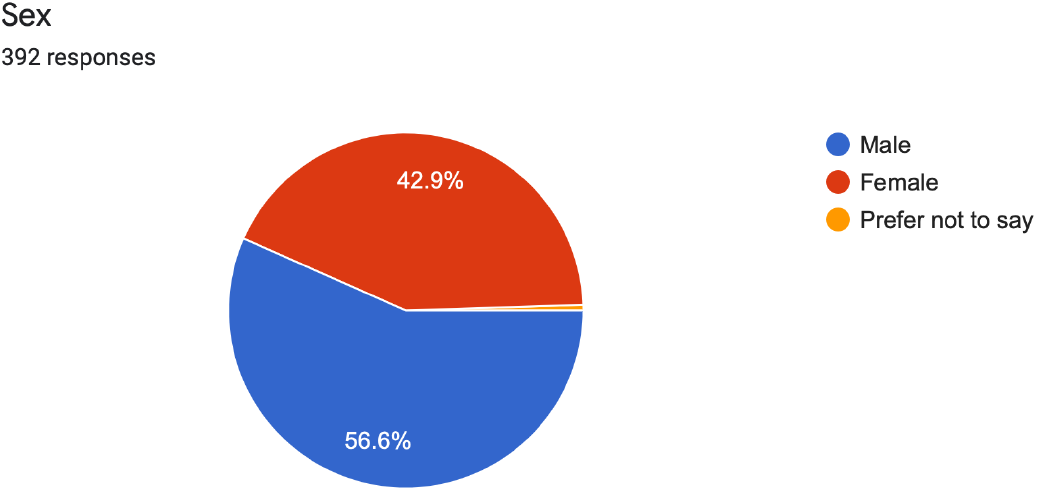

### 2. FAMILY INCOME

Majority (78.8%) of the participants belonged to Class I Socioeconomic Status as per Modified BG Prasad Classification for the year 2020. (We used 2020 scale as the peak of the pandemic and lockdown was in the year 2020)

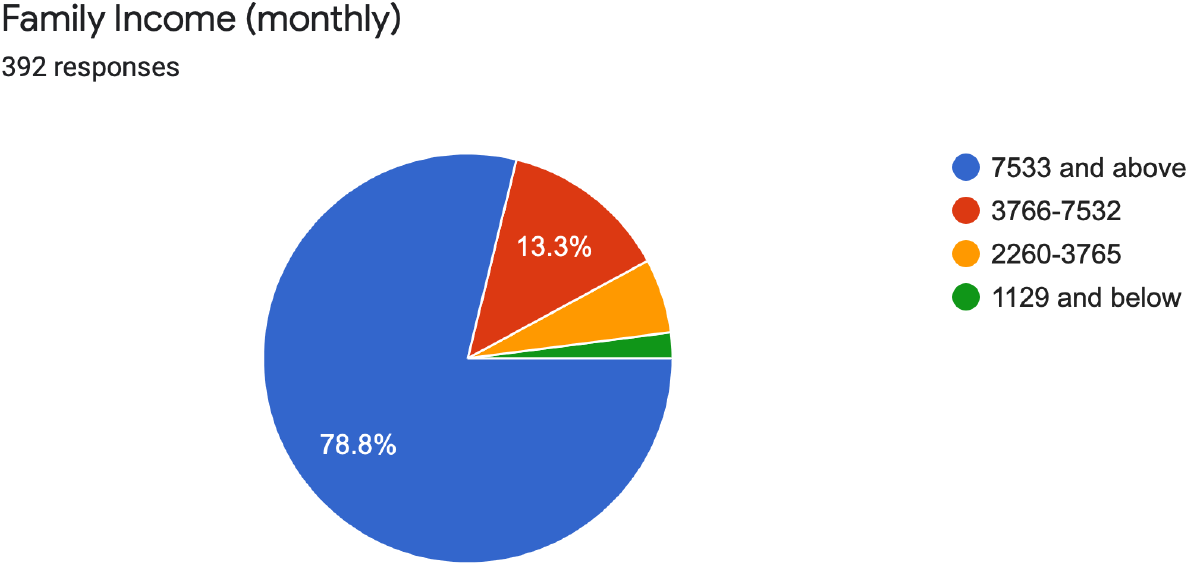

### 3:- AREA OF RESIDENCE

Majority (67.6%) of the respondents were urban residents.

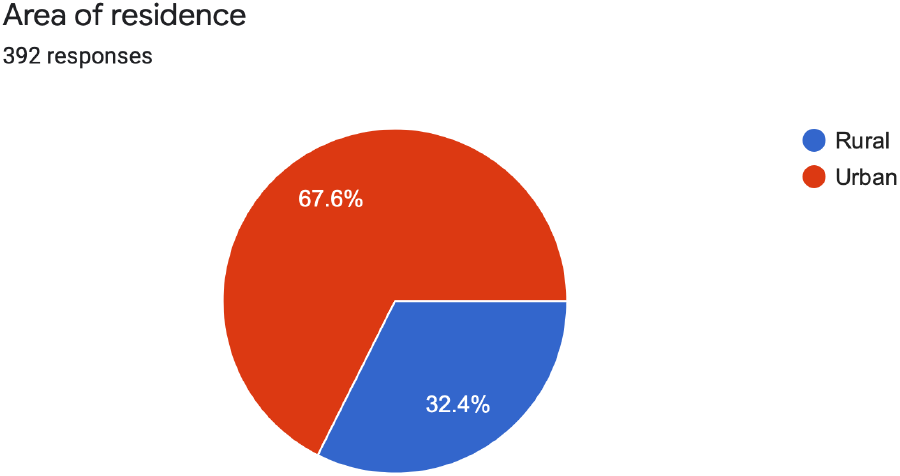

### 4:- TYPE OF FAMILY

Majority (80.1%) of the participants were from nuclear families.

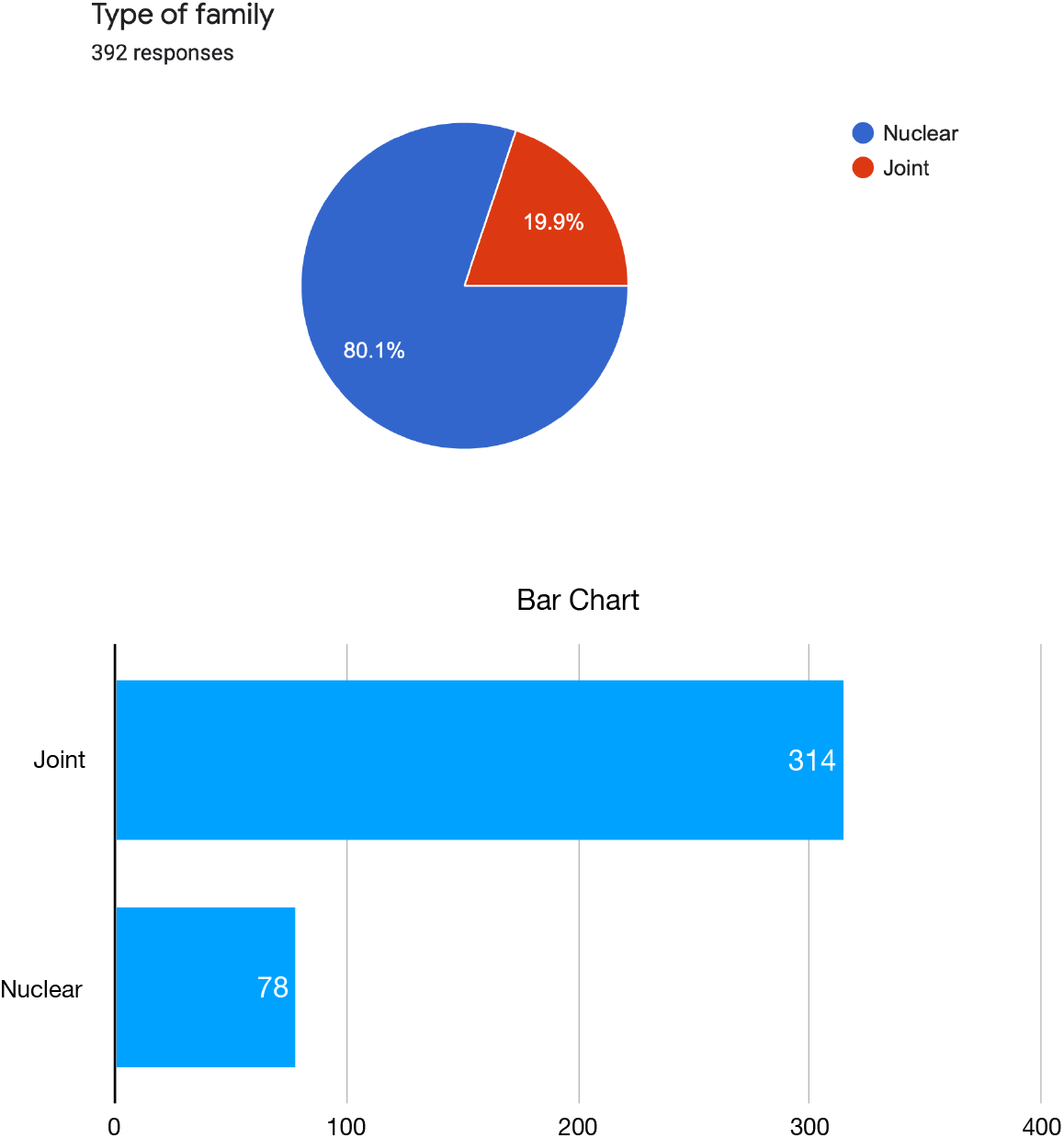

### 5:- ANY CASE OF JOB LOSS

13.8% of the respondents reported loss of job in their family.

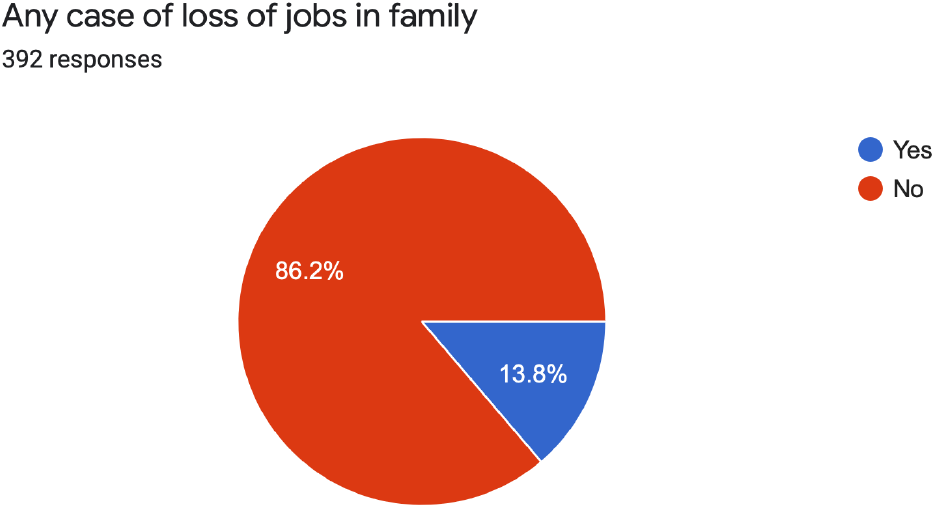

## ANXIETY SCALE INTERPRETATIONS

Based on the Zung self rating Anxiety scale

1: 7.4% of the respondents said they felt afraid without any reason at all, on the other hand the majority (41.1%) reported to have it a little of the time.

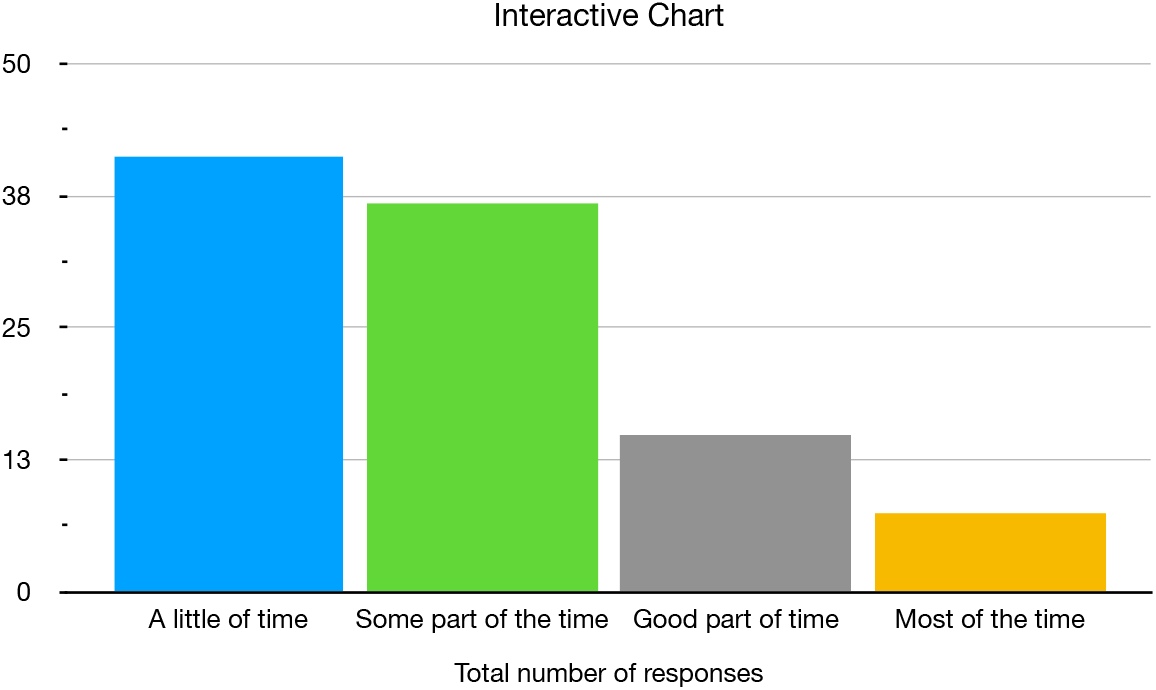

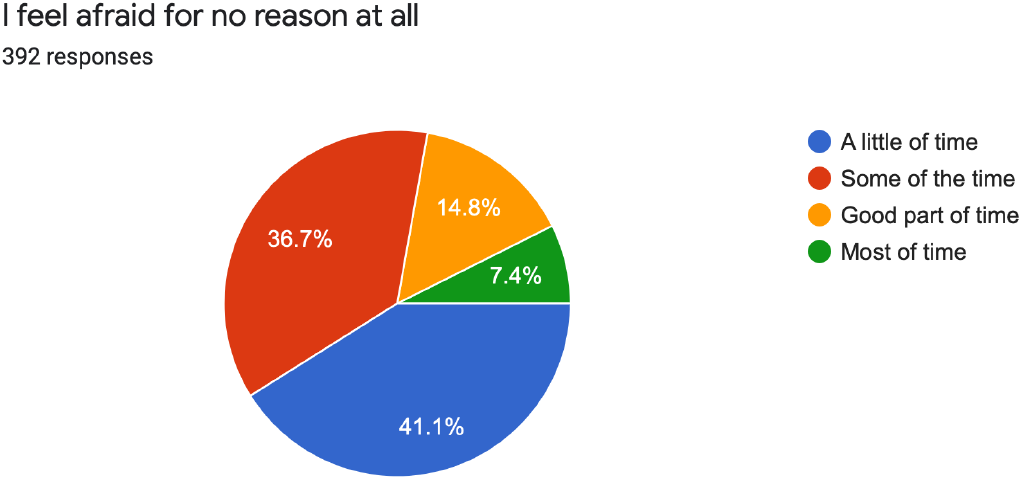

2: 12.5% of the participants told they felt more nervous and anxious than usual while the majority of 38.5% reported to have it for a very little of the time. It should be noted that 32.9% said to have it sometimes.

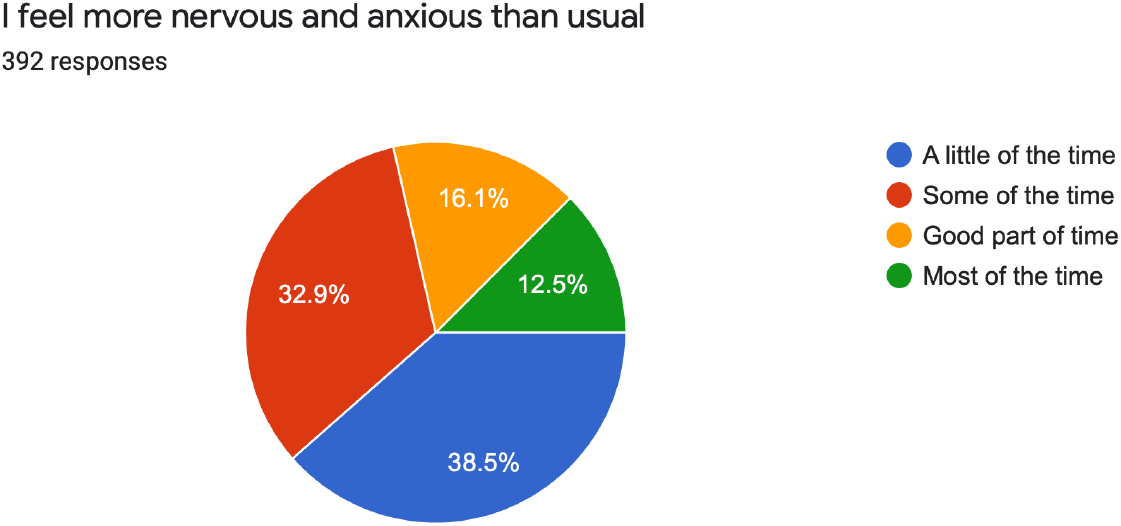

3: 18.1% of the participants said they get upset or feel panicky for most of the time. It should be noted that 21.4% and 28.8% of the participants felt the same for good part of time and some part of time respectively.

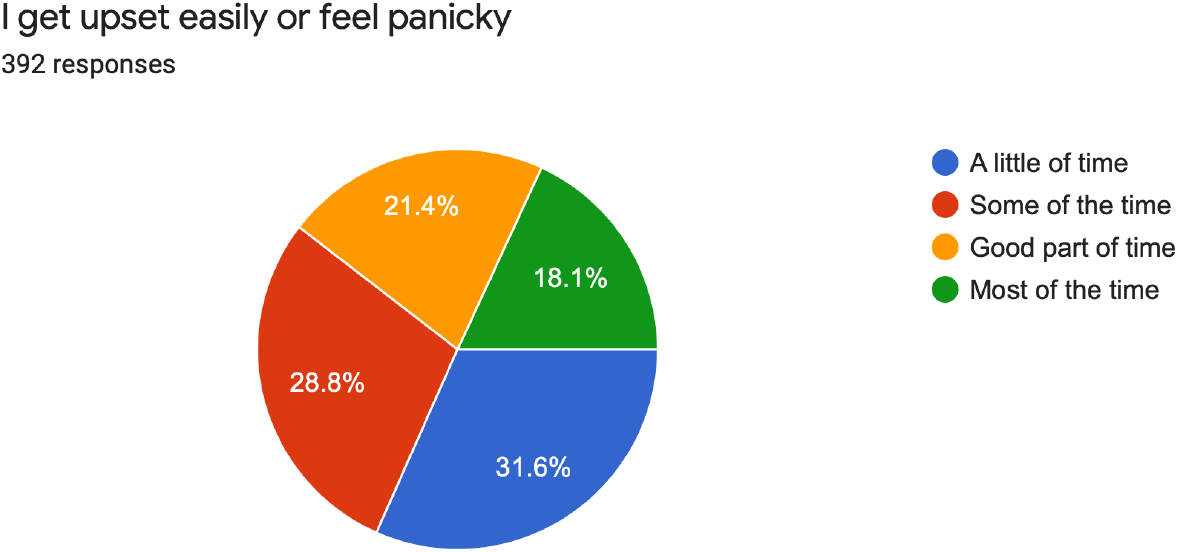

4: 12.2% of the respondents said that they feel like falling apart and going into pieces. We also found that 30.4% and 14.3% said that they feel the same for some of the time and good part of time respectively.

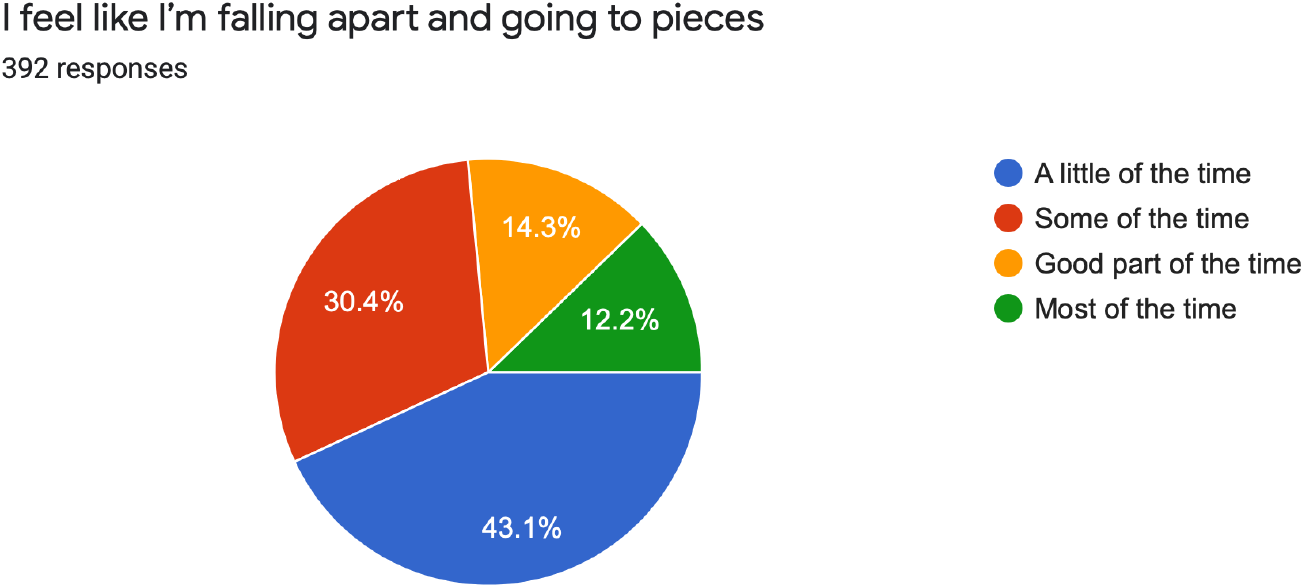

5: 29.8% of the respondents said that most of the time they don’t feel everything is all right and something bad can happen. On the other hand 17.3% of the candidates said that this happens most of the time, while 21.9% of the candidates said this happens for good part of time.

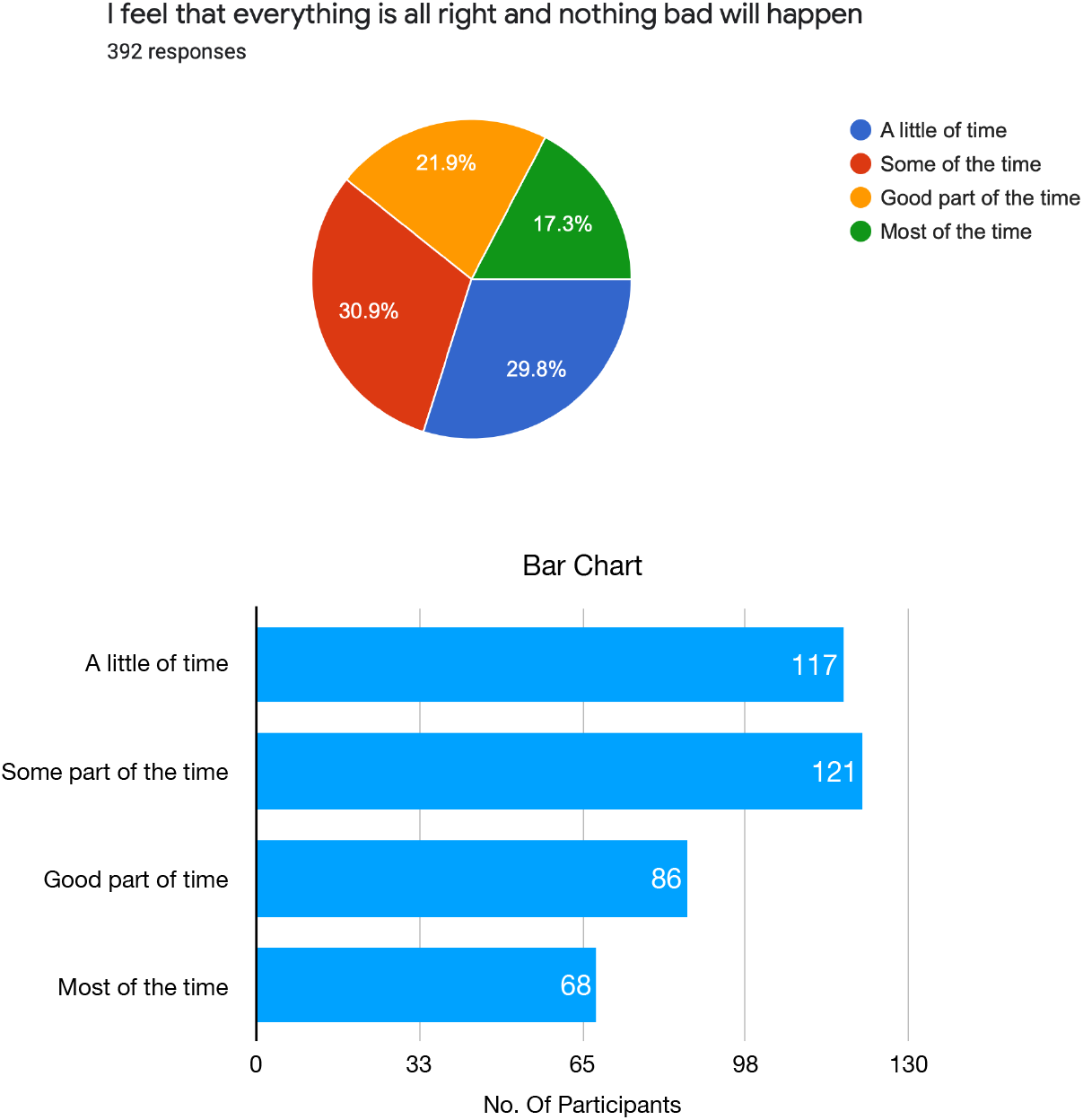

6: 13(3.3%) and 17(4.3%) candidates said that their arms and legs tremble for most of the time and good part of time respectively. Also 45 candidates (11.5%) reported the same for some part of the time.

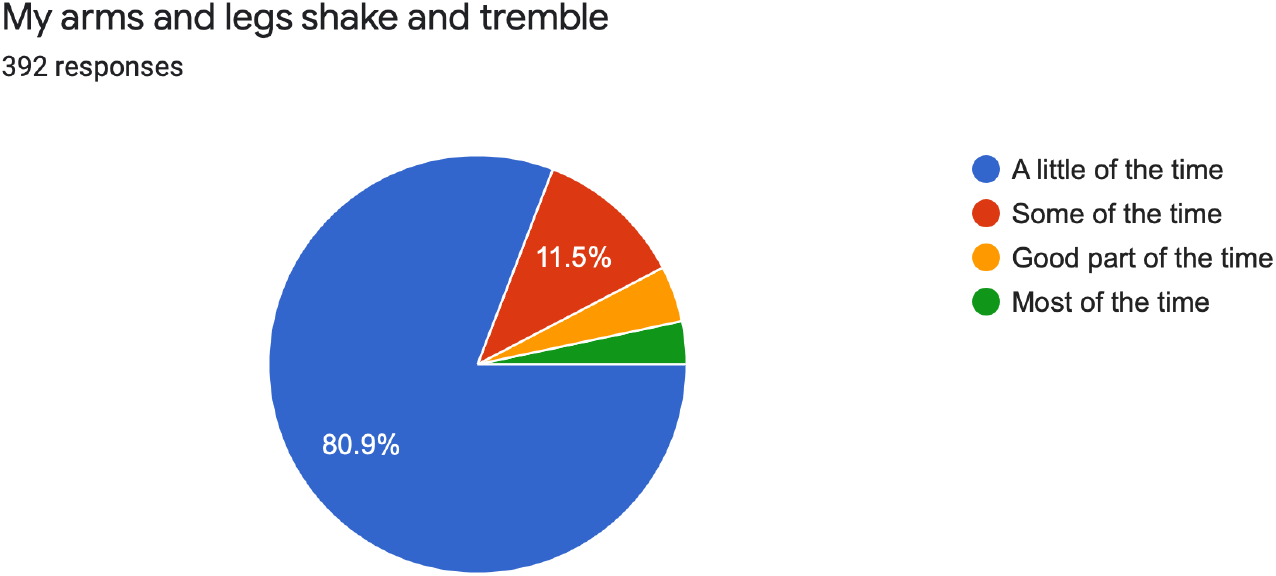

7. 9.9% and 16.1% of the candidates reported that they are bothered by headaches, neck and back pain for most of the time and good part of time respectively. On the other hand, 26.5% of the candidates reported the same for some part of the time.

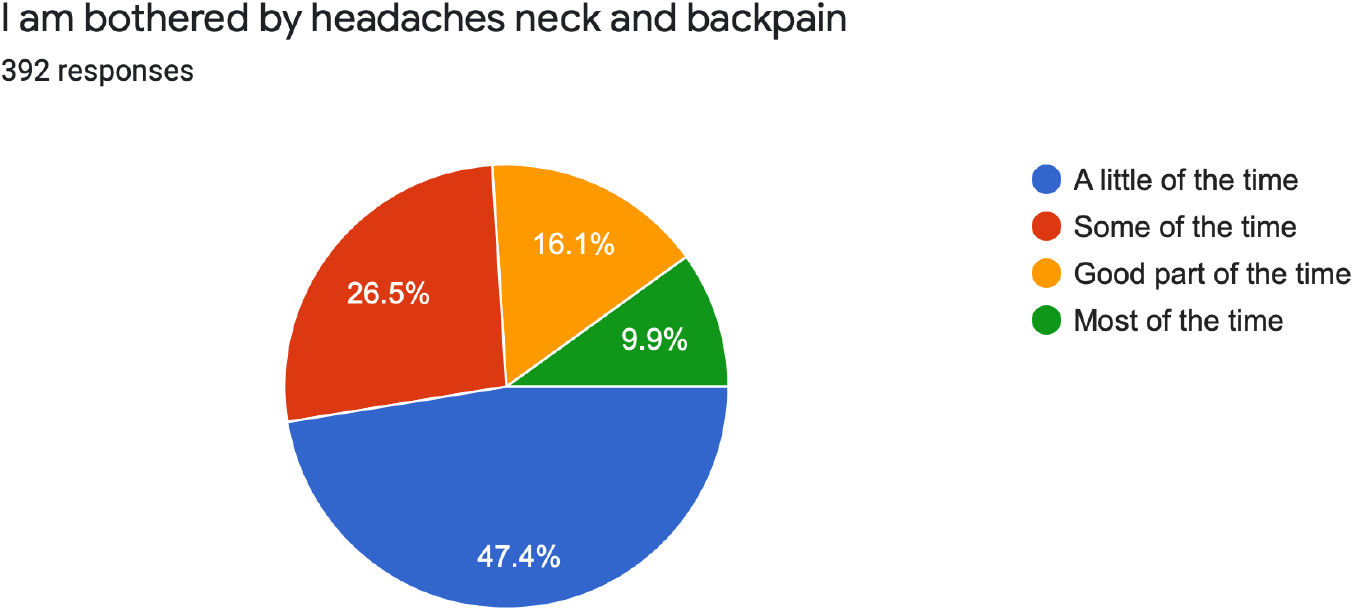

8. 27.8% of the candidates said that they feel calm and can sit still easily for only a little part of the time. While 29.3% are able to do so for only some part of the time.

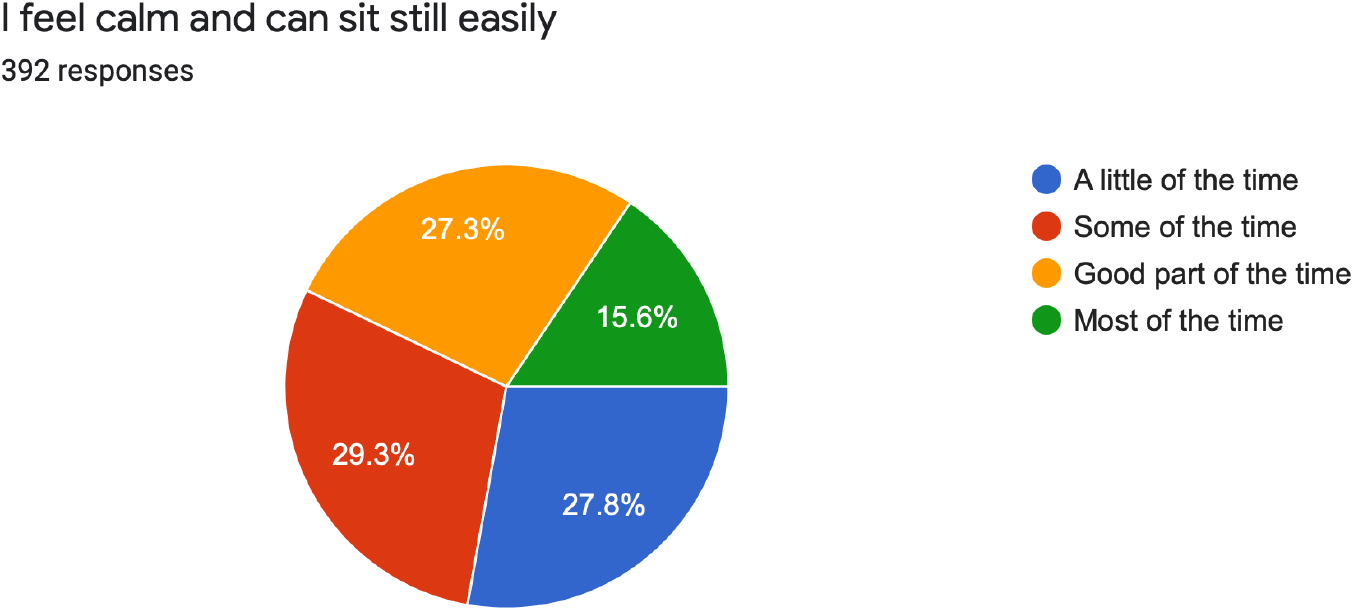

9. 16.1% of the candidates felt weak and get tired easily for most part of the time. On the other hand 17.9% of the candidates did the same for a good part of the time.

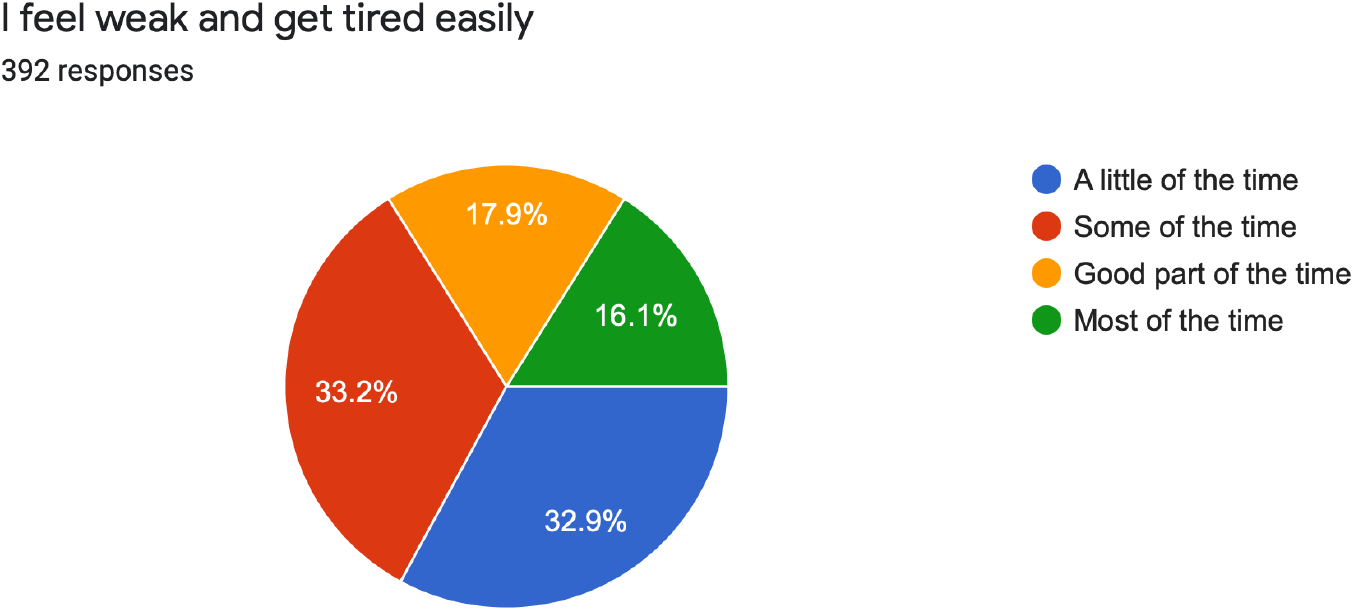

10. 17 (4.3%) candidates reported that they can feel their heart beating fast for most of the time. On the other hand, 12.5% of the candidates felt the same for good part of the time.

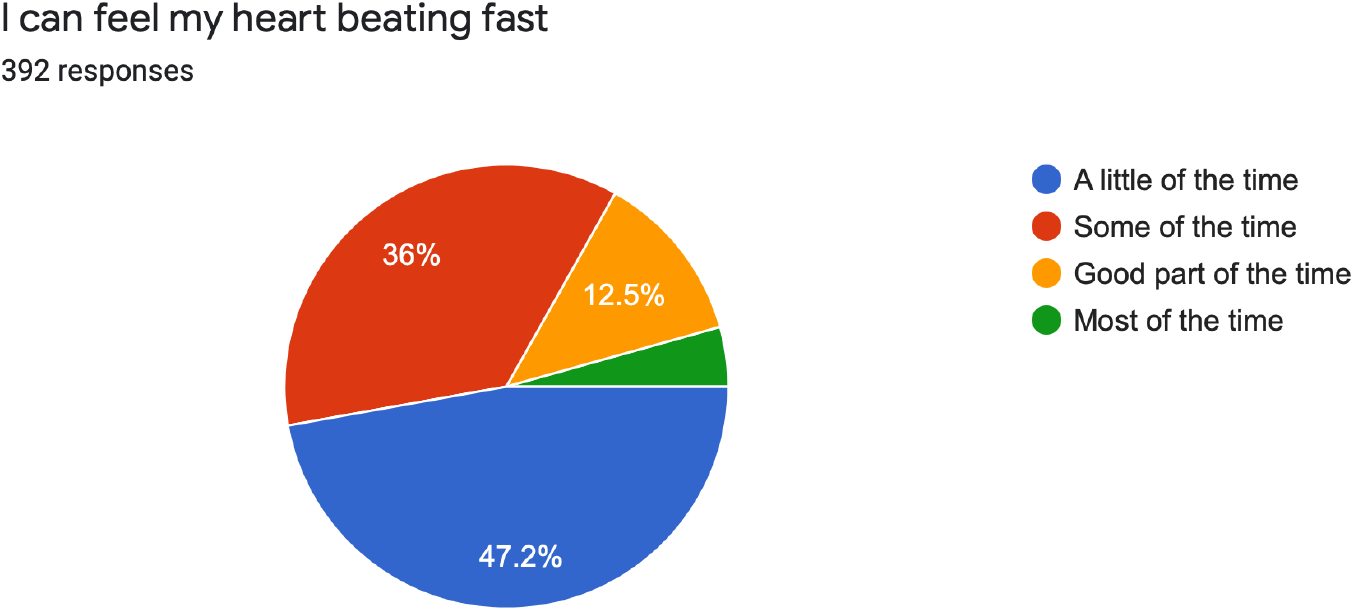

11. 14(3.6%) of the respondents reported that they are bothered by dizzy spells for most of the time. On the other hand 27 candidates (6.9%) said they felt the same for good part of the time.

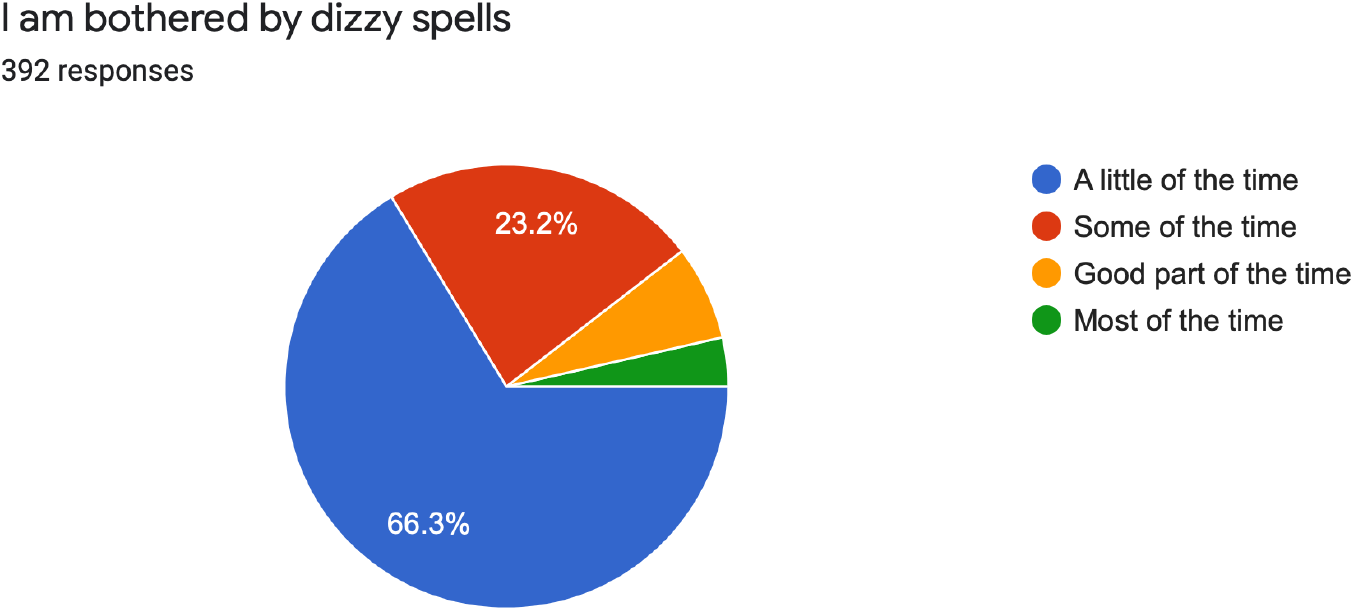

12. 8.9% of the respondents said that they can breadth in and out for only a little part of the time while 6.4% of the candidates reported the same for some part of the time.

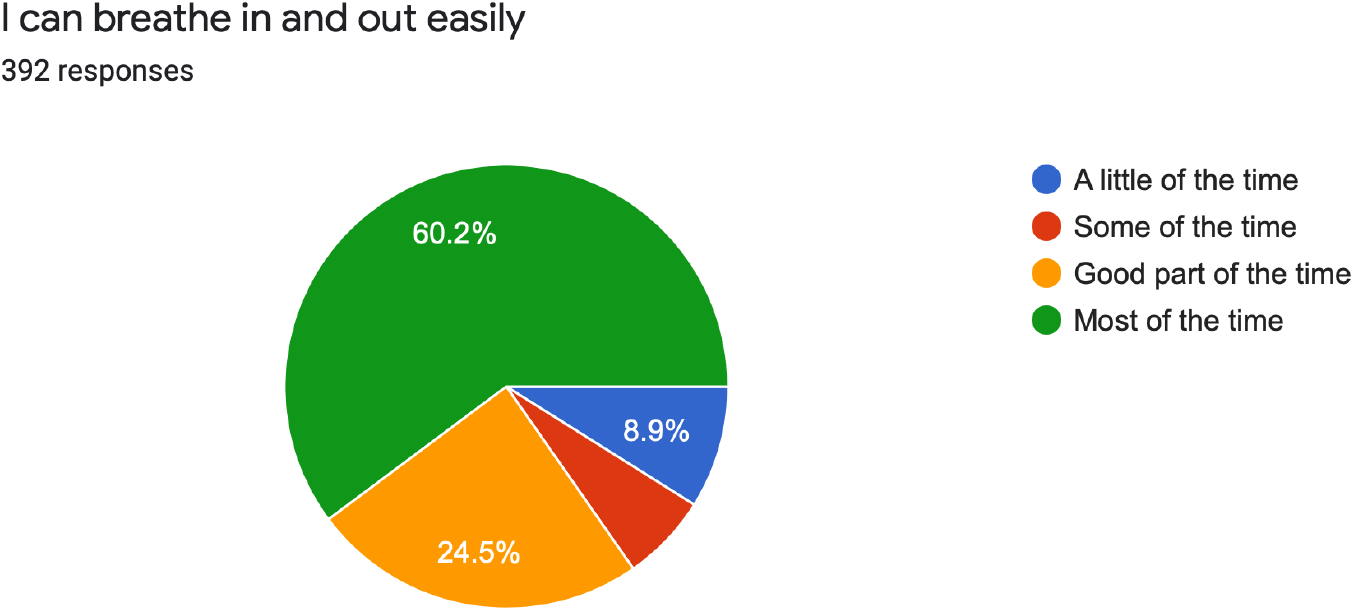

13. 9 (2.3%) of the candidates said they had fainting spells or feel like it fo most of the time. While 2.8% of the candidates felt the same for good part of the time.

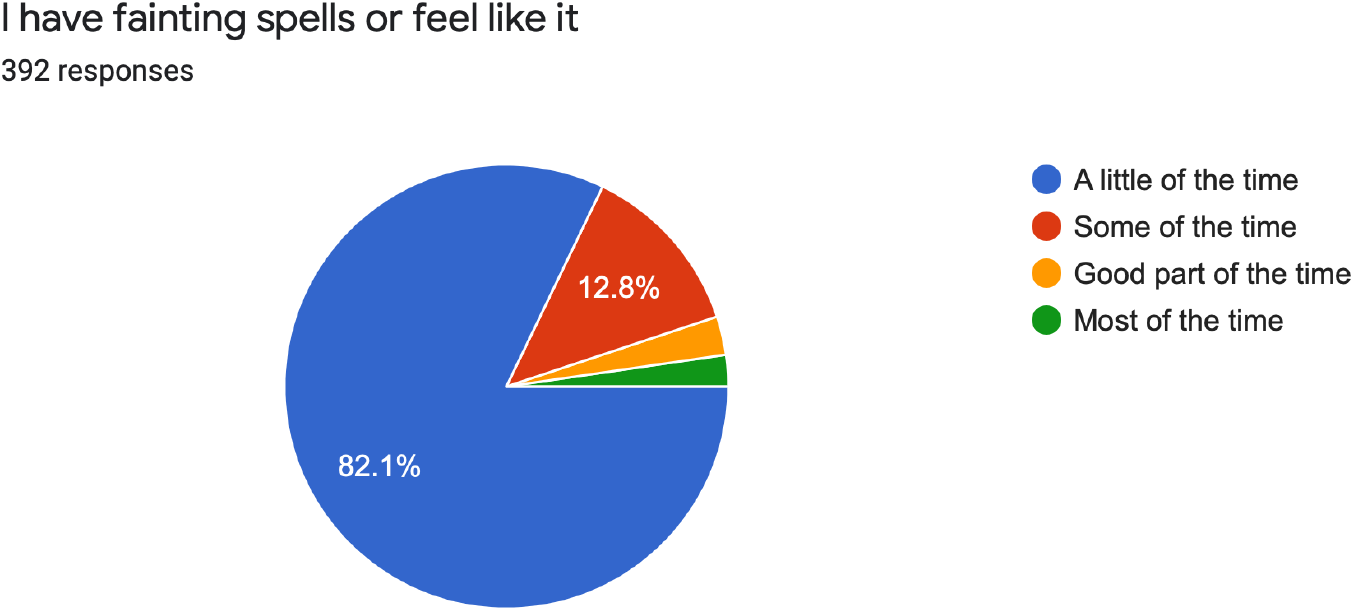

14. 13 (3.3%) of the candidates said that they felt numbness and tingling in their fingers and toes for most of the time while 21 (5.4%) of the candidates reported the same for good part of time.

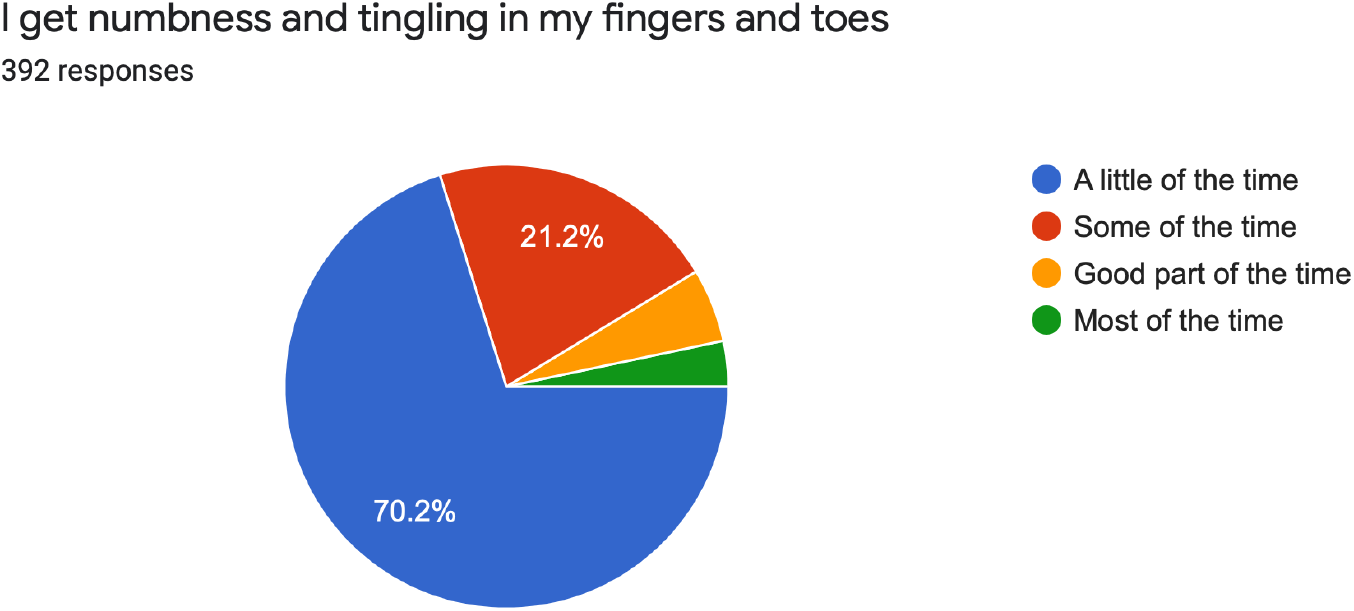

15. 8.2% of the respondents are bothered by stomach or indigestion while 10.2% of the candidates are bothered by the same for good part of the time.

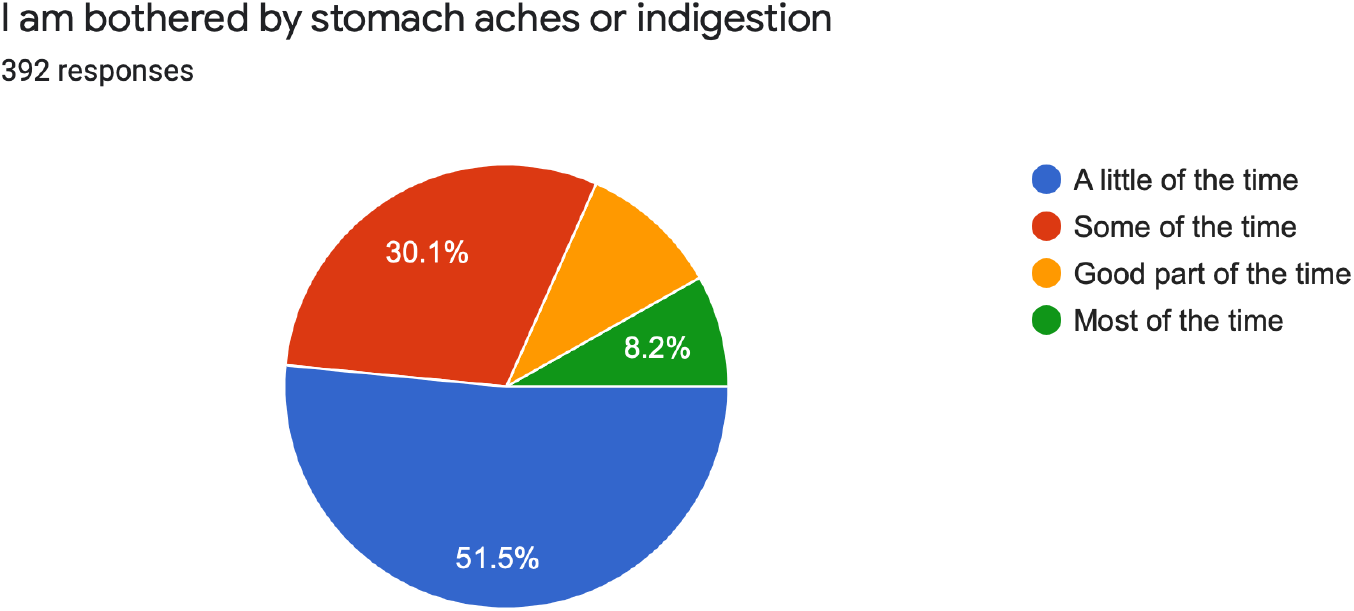

16. 15 (3.8%) of the respondents reported that they have to empty bladder often for most of the time while 12.2% of the respondent did the same for good part of the time.

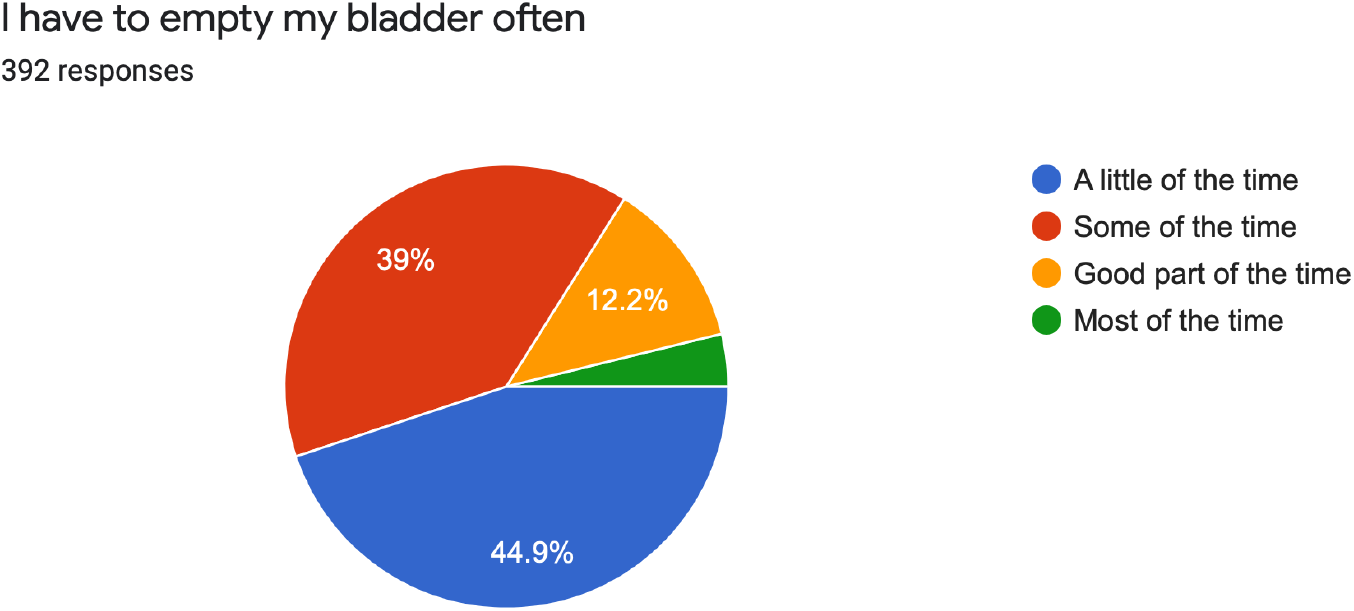

17. 22.2% of the candidates reported to have usually dry and warm hands for most part of the time while 16.8% of the candidates reported the same for good part of the time.

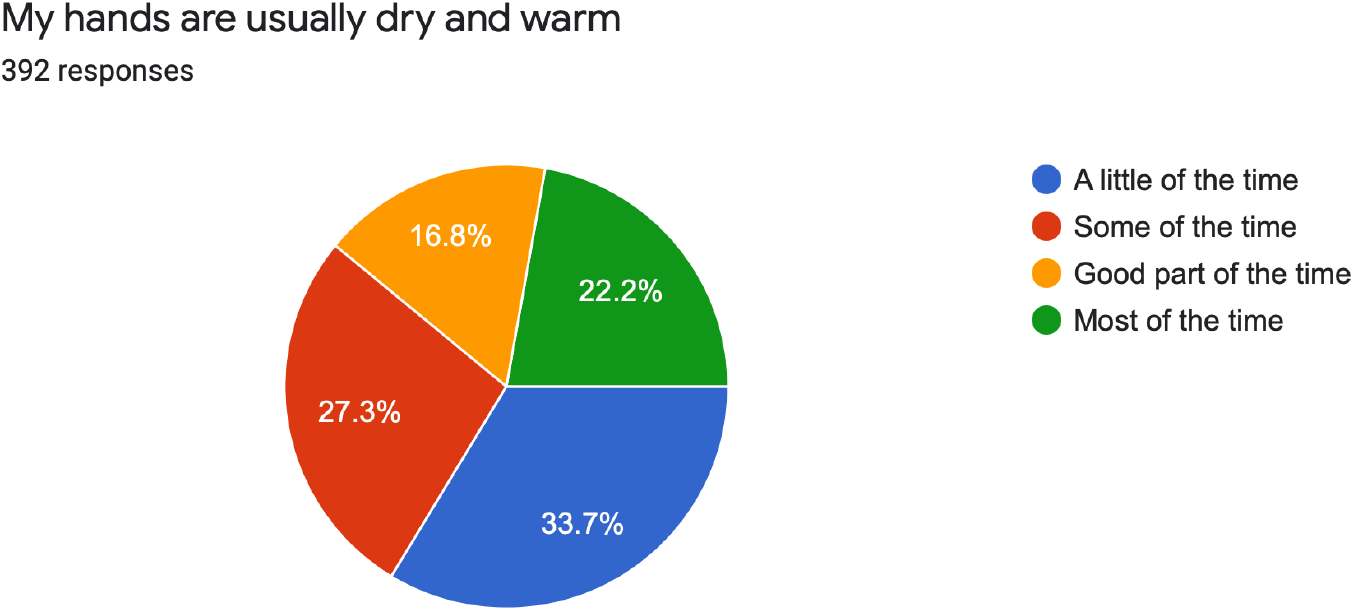

18. 12(3.1%) of the candidates said their face got hot and blushes for most of the time while 9.7% of the candidates felt the same for good part of the time.

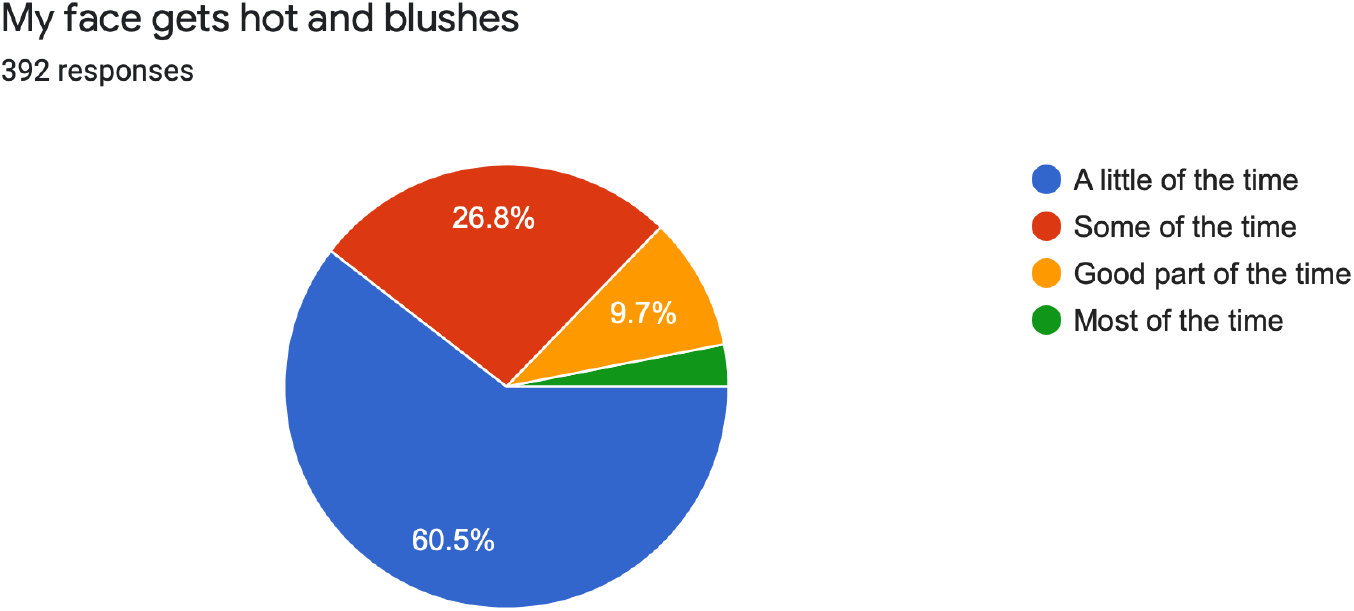

19. 27.8% of the candidates said they fell asleep and get good night’s rest for only little of the time. While 24.2% of the of the respondents had the same for only some of the time.

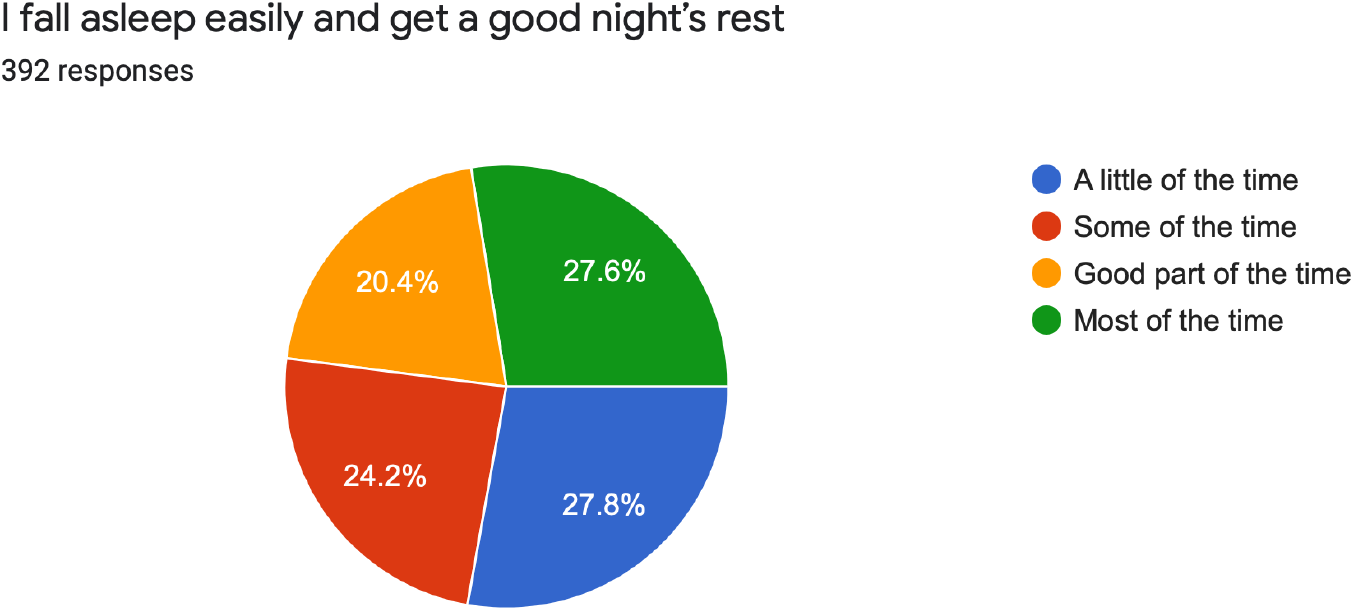

20. 8.4% of the respondents had nightmares for most of the time while 9.2% of the candidates had the same for good part of the time.

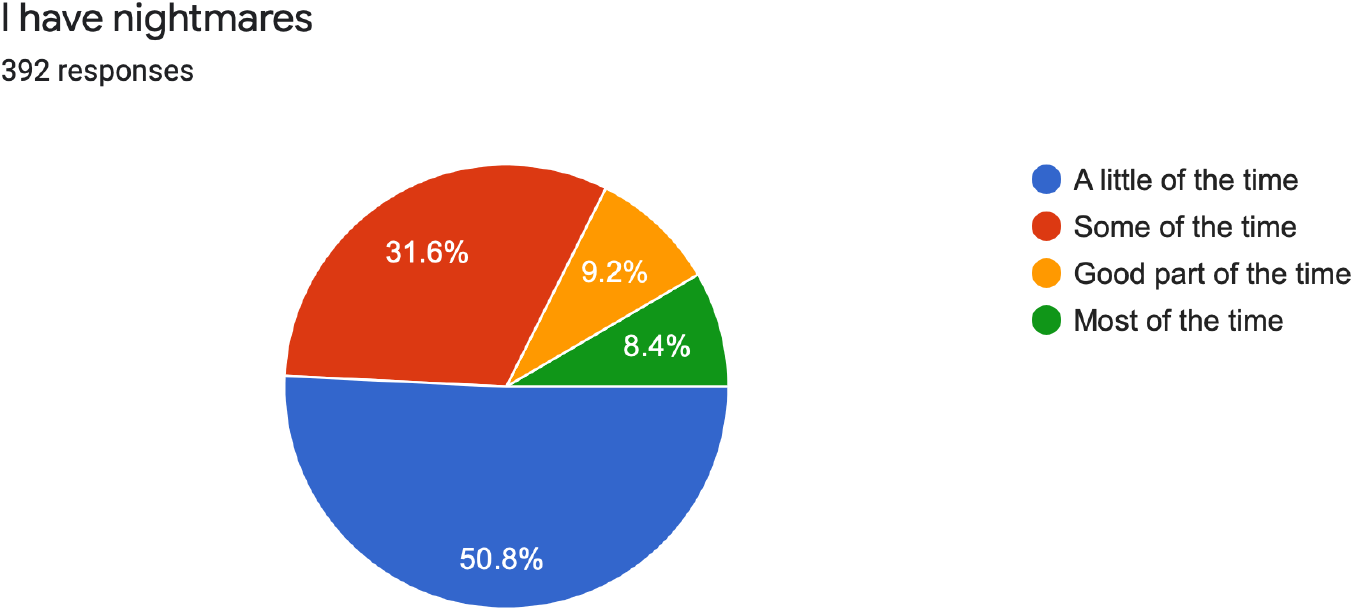

## DEPRESSION SCALE INTERPRETATIONS

Based on the Zung self rating Depression scale

21. 9.9% of the candidates felt down hearted and blue for most of the time while 16.1% of the candidates felt the same for good part of the time.

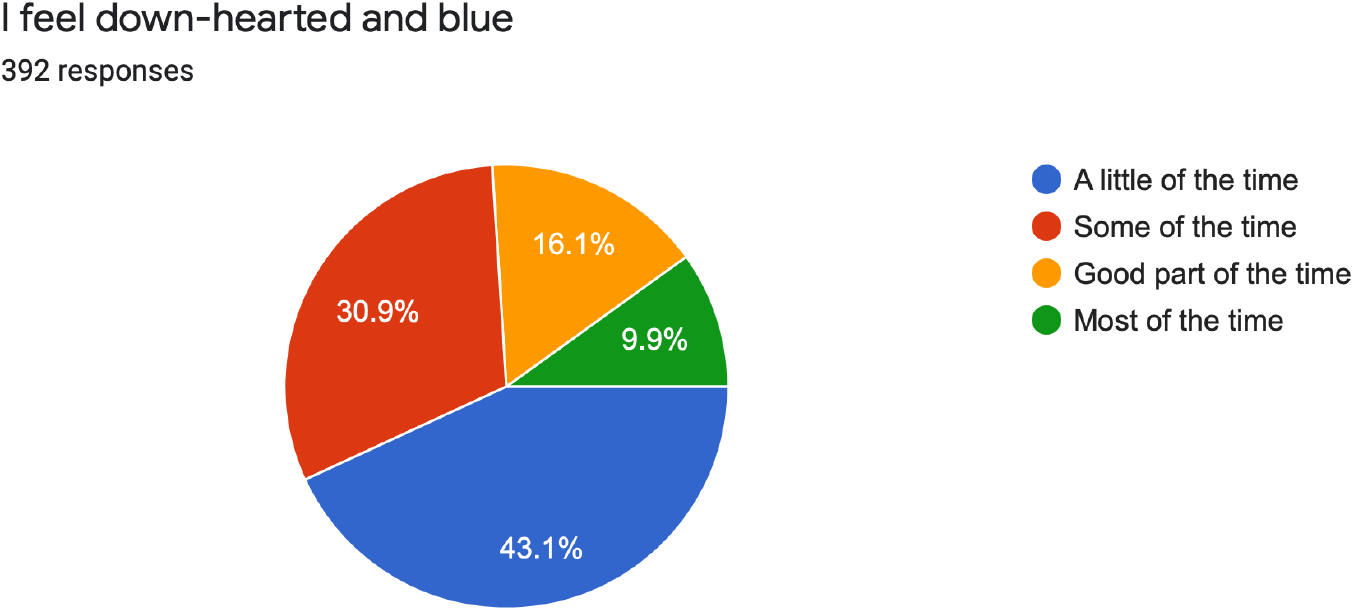

22. 23.7% of the respondents said they feel morning is the time when they feel the best for a little of the time. While 25.8% of the candidates felt the same for some part of the time.

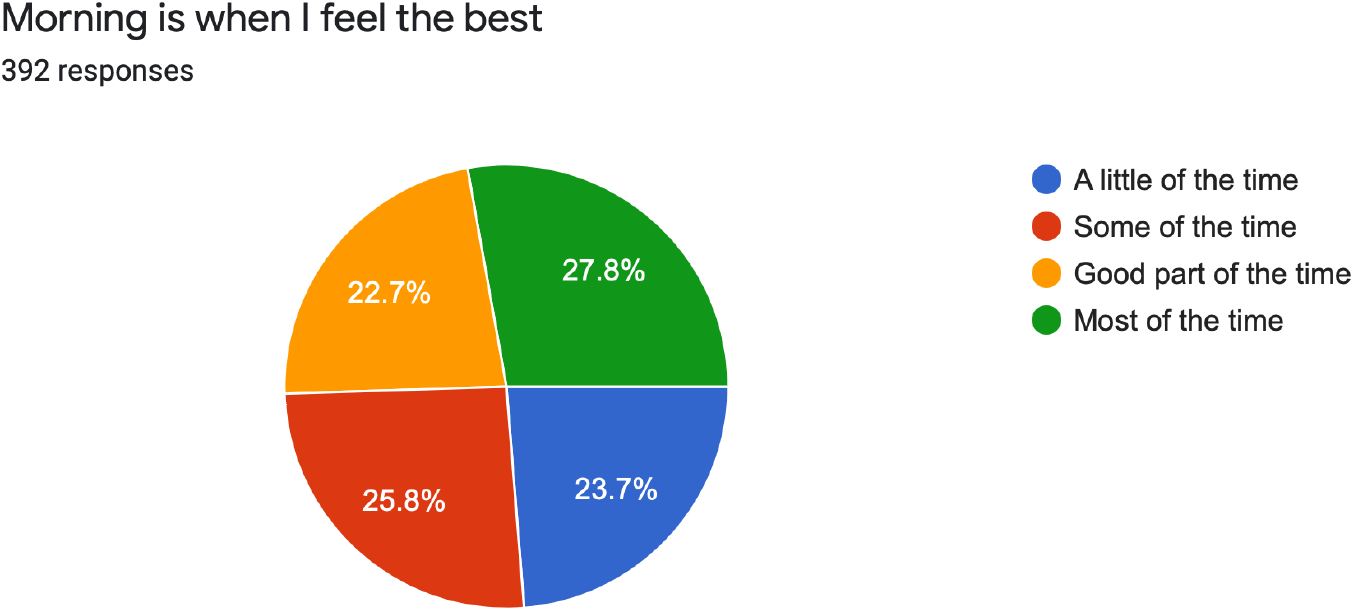

23. 10.7% of the candidates said they had crying spells of feel like that for most of the time while 14.5% of the candidates felt the same for good part of the time.

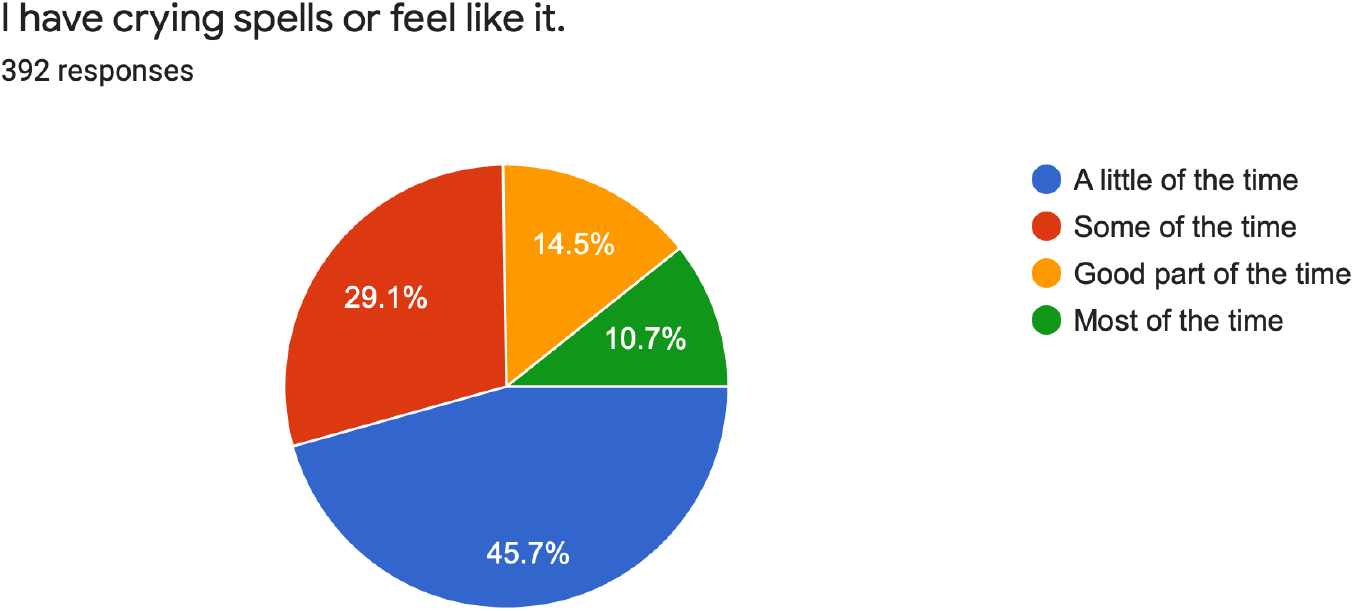

24. 19.9% of the candidates had trouble in sleeping at night for most of the time, while 13.5% of the candidates had the same trouble good part of the time.

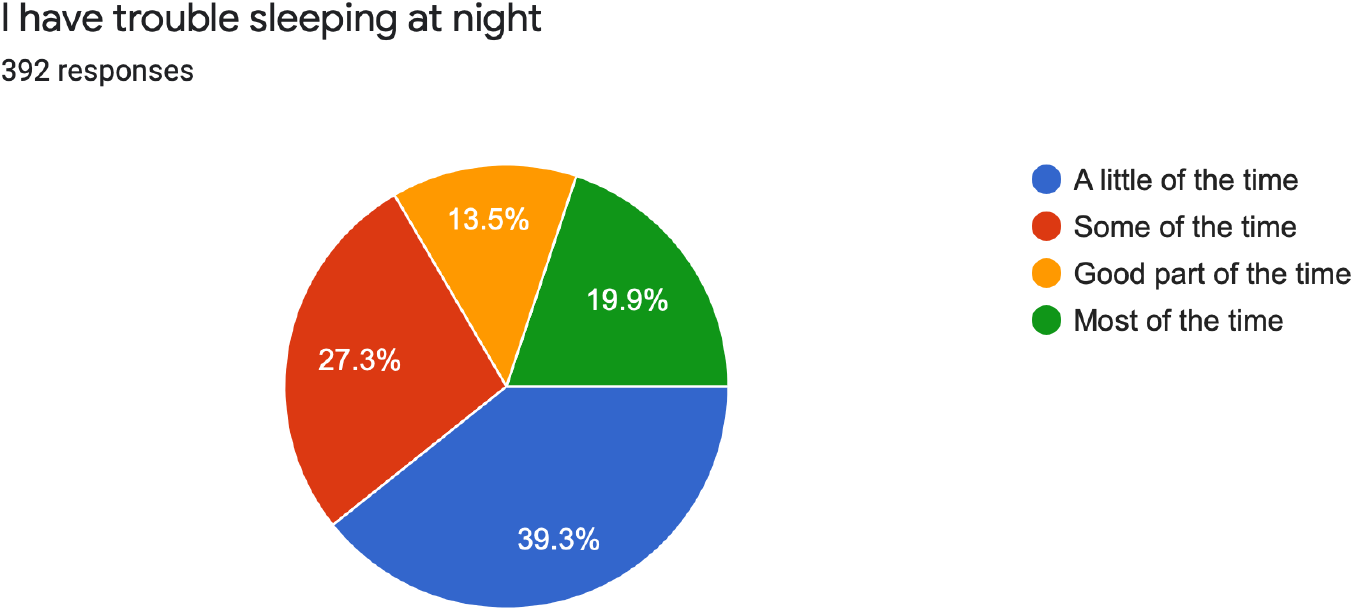

25. 18.1% of the participants reported that they ate as much as they used to for a little of the time while 25.5% of the participants ate as much as they used to for some of the time.

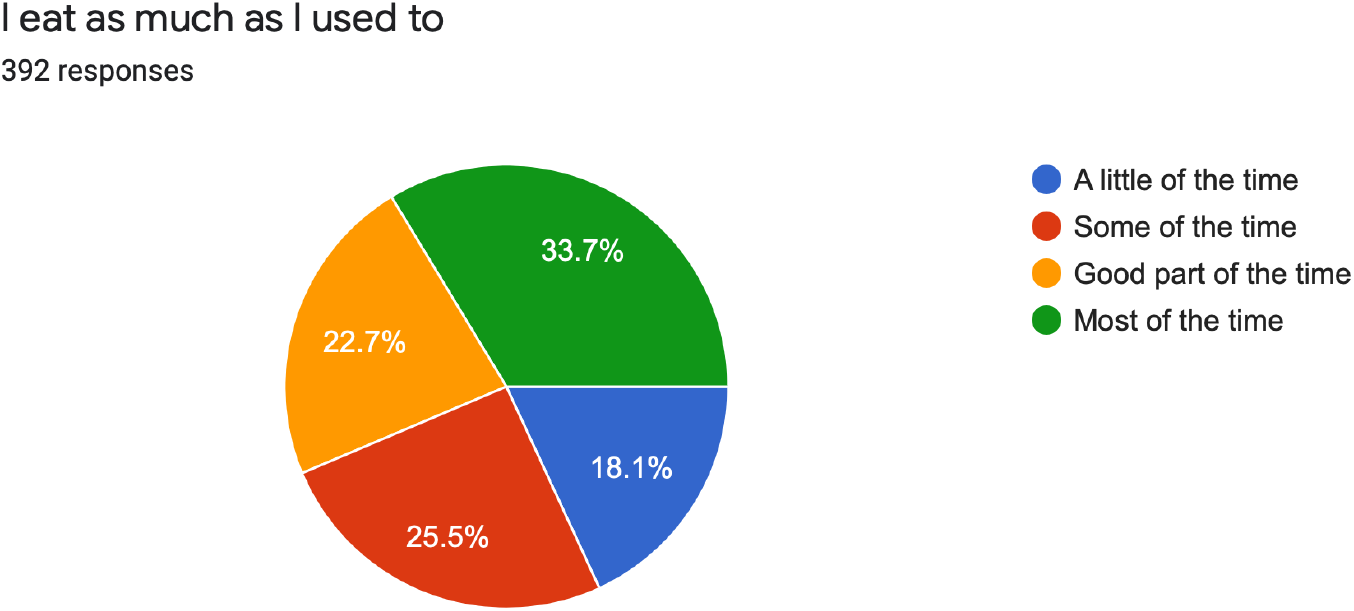

26. 41.1% of the participants still enjoyed sexual activity for little of the time. On the other hand 22.2% for the participants enjoyed the same for some part of the time.

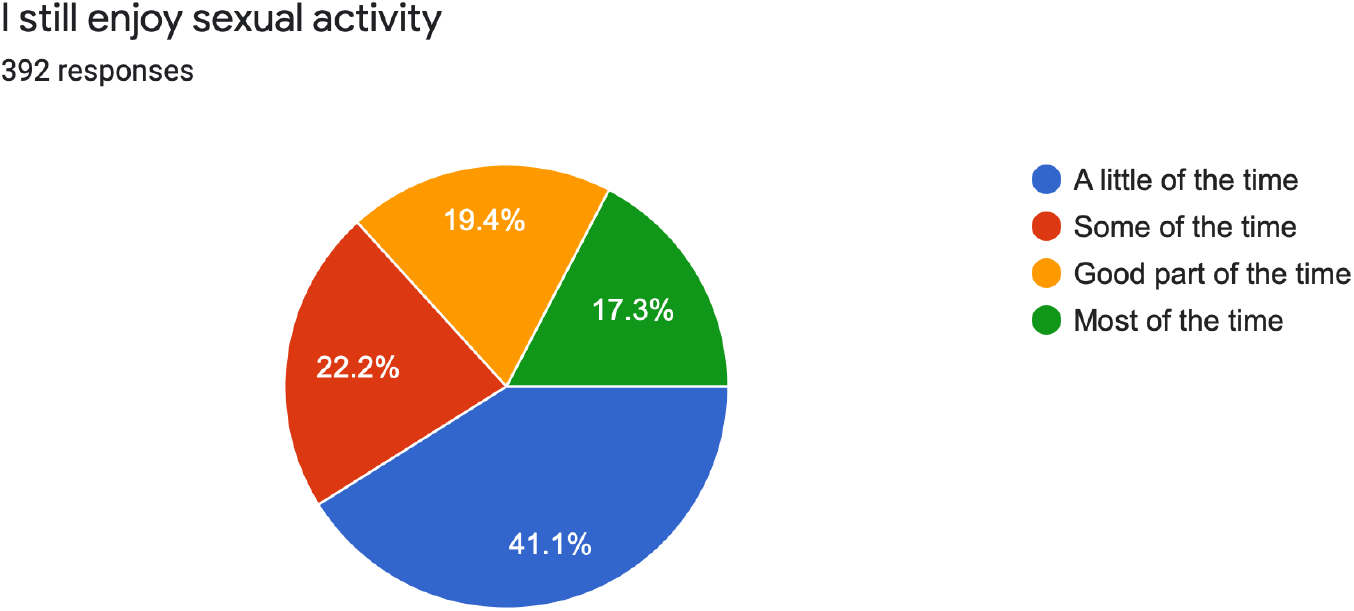

27. 7.1% of the candidates reported that they are losing weight for most of the time while 8.4% of the candidates reported the same for good part of time

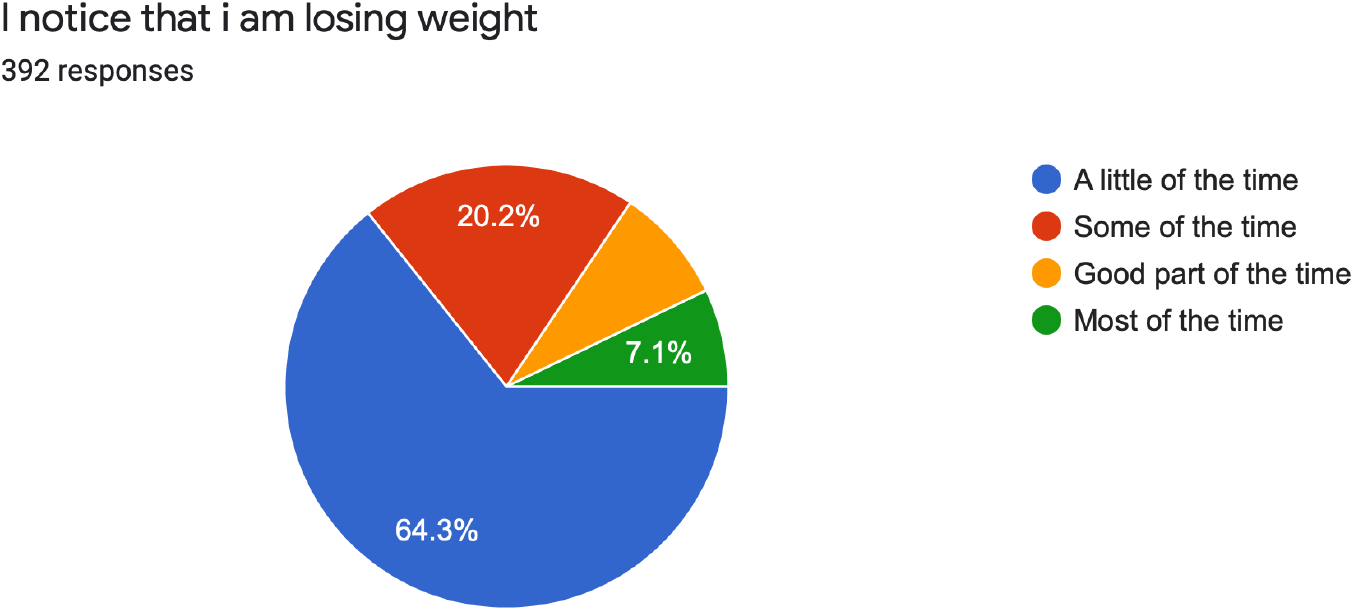

28. 27 (6.9%) of the candidates reported to have trouble with constipation for most of the time. On the other hand 29 (7.4%) of the candidates reported the same trouble for good part of time.

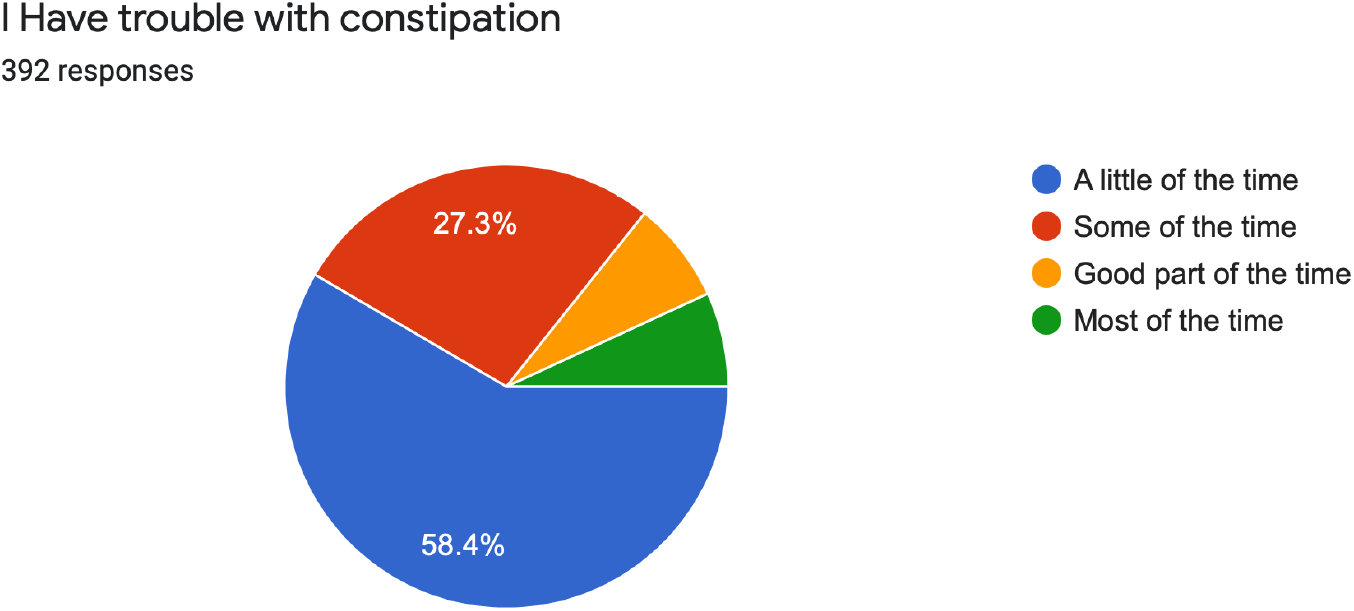

29. 21 (5.4%) of the respondents said their heart beats fasted than usual for most of the time while 8.9% of the respondents felt the same for good part of the time.

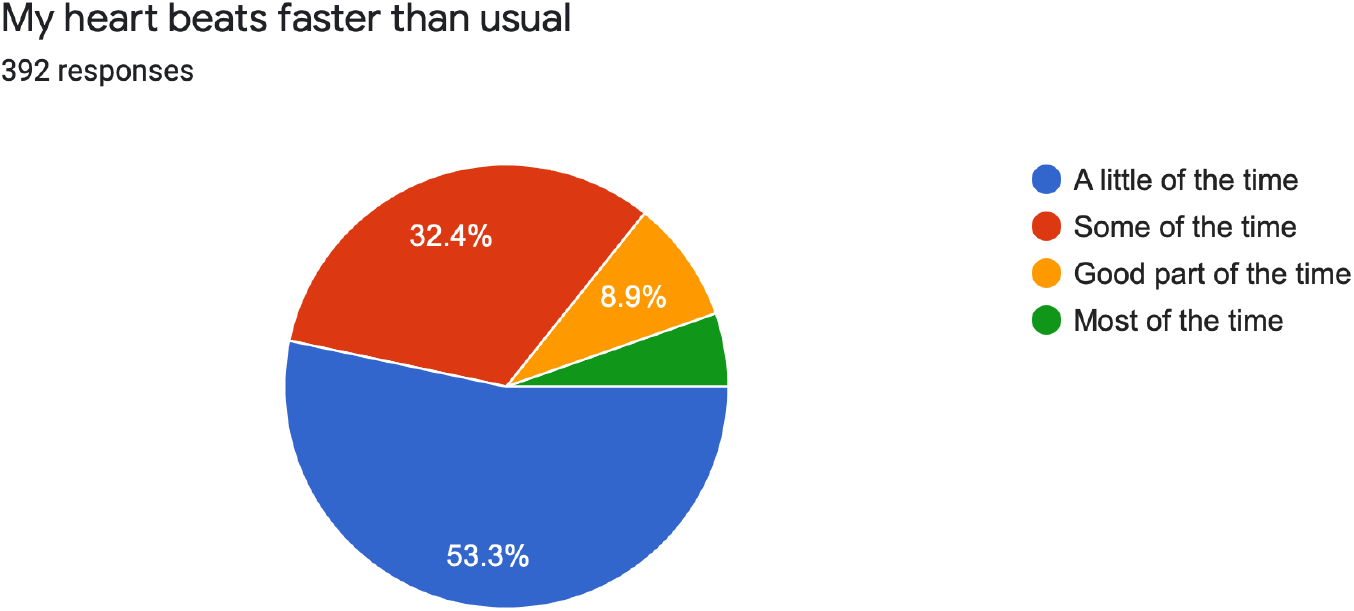

30. 16.1% of the respondents said that they get tired for no reason for most of the time. On the other hand 21.4% of the respondents had the same issue for good part of the time.

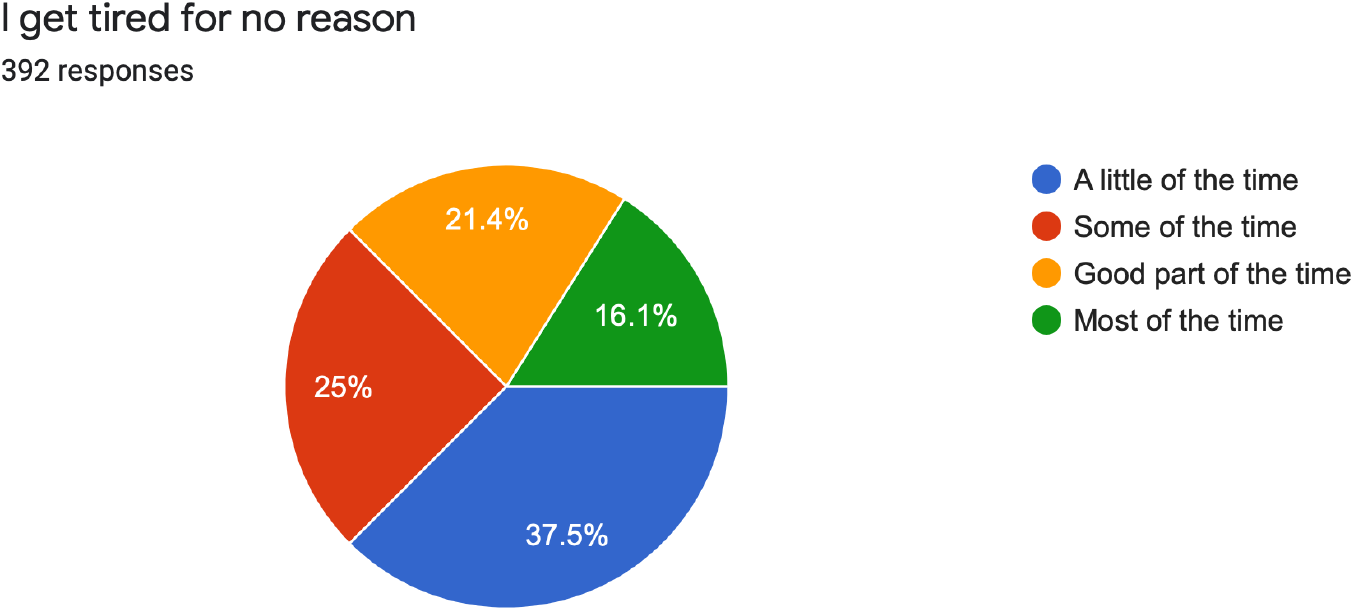

31. 36% of the candidates said their mind was as clear as it used to be for a little of the time.

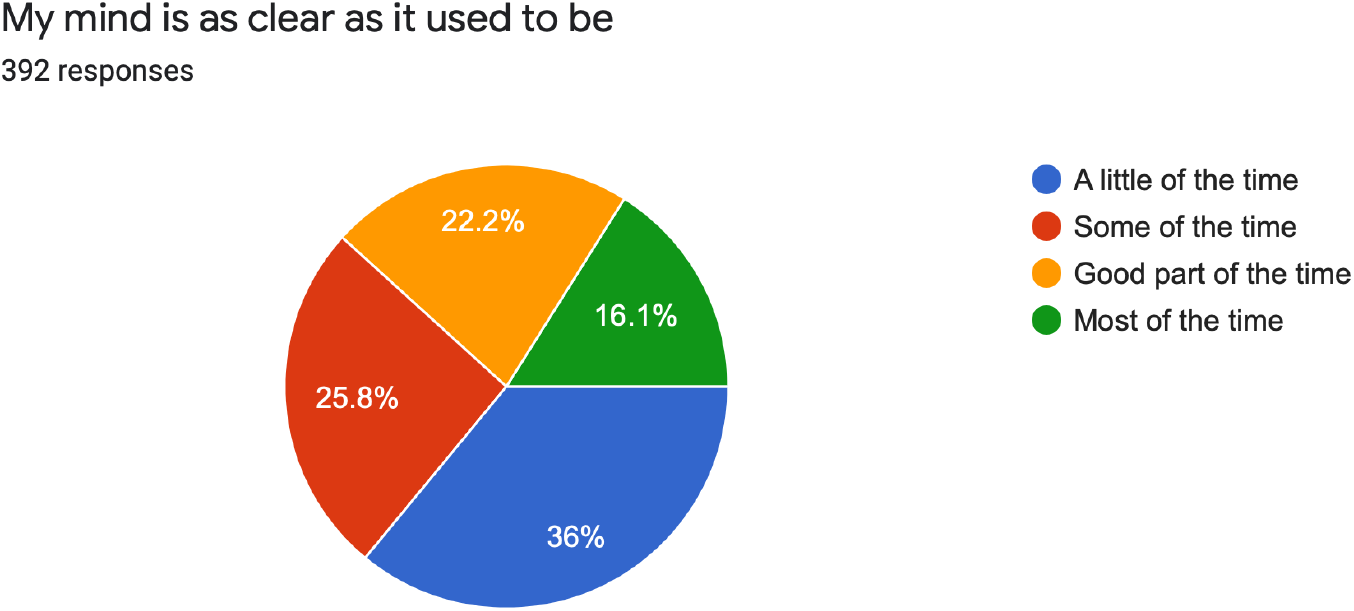

32. 31.9% of the respondents were found to do things easily as the used to only for a little of the time while 29.1% of the respondents felt the same for some part of the time.

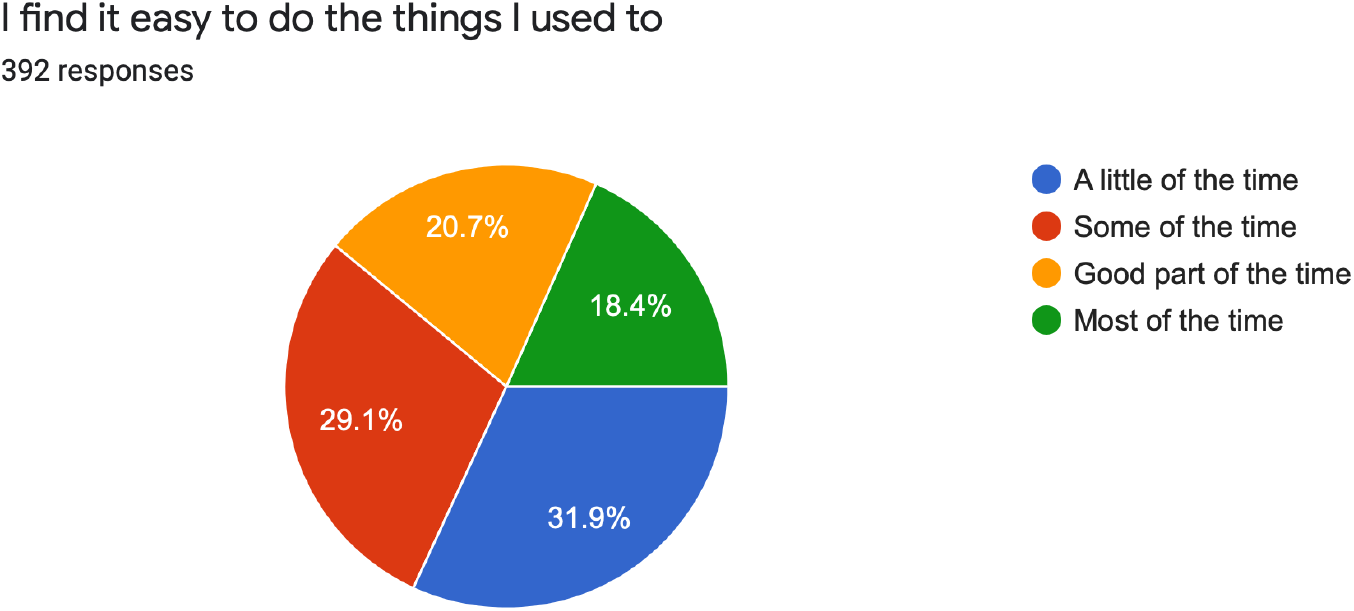

33. 11.2% of the candidates felt they are restless and can’t keep still for most of the time, while 17.1% of the candidates felt the same for good part of time.

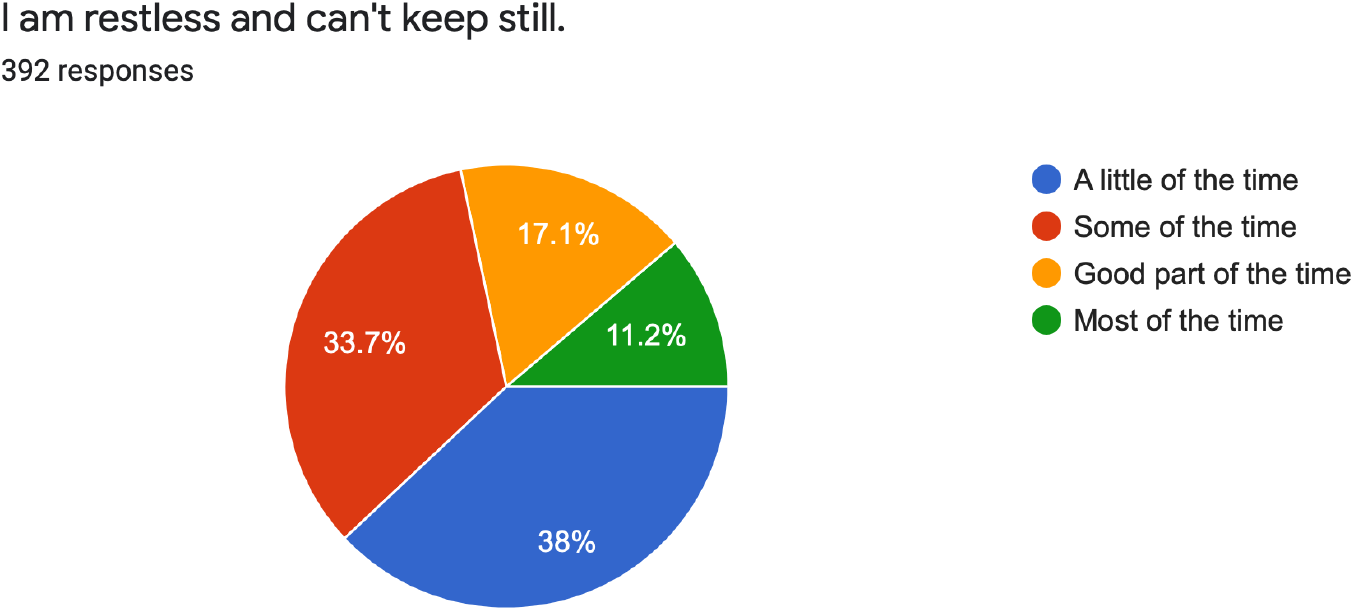

34. 27% of the respondents said they felt hopeful about the future only for a little of the time, while 27.6% of the respondents felt the same for some of the time.

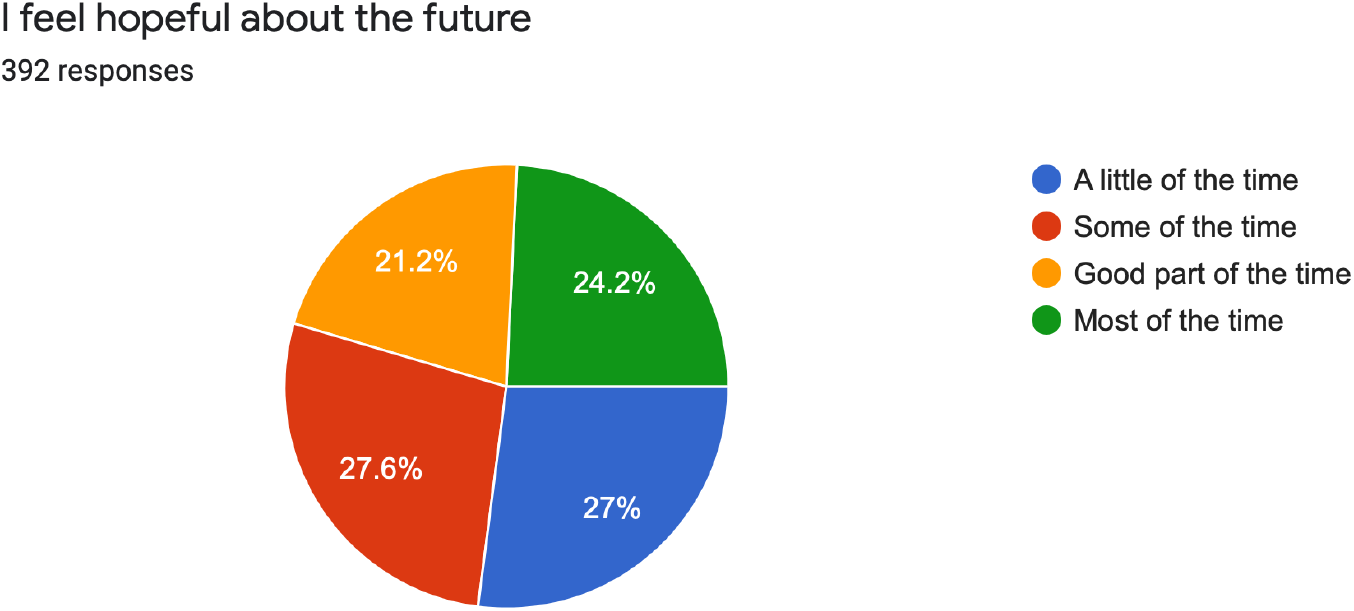

35. 19.9% of the candidates felt they were more irritable than usual for most of the time while 19.6% of the candidates felt the same for good part of the time.

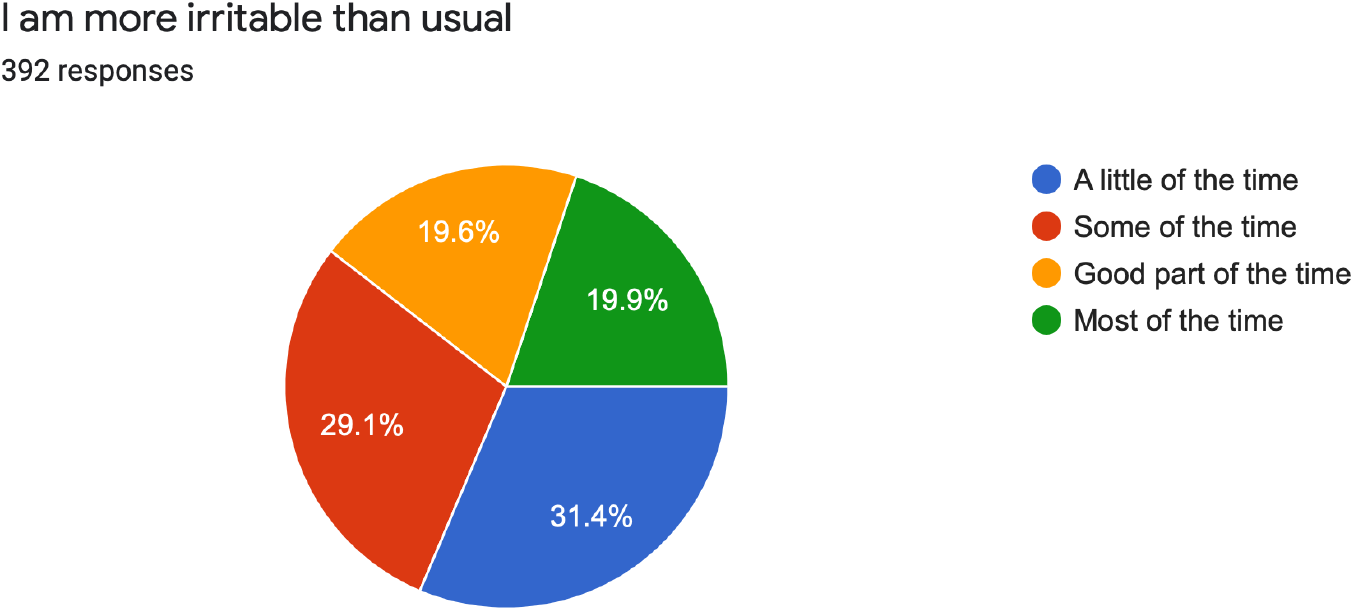

36. 32.7% of the respondents found it easy to make decisions only for a little of the time while 36.5% of the respondents found that it was easy to make decisions only for some of the time.

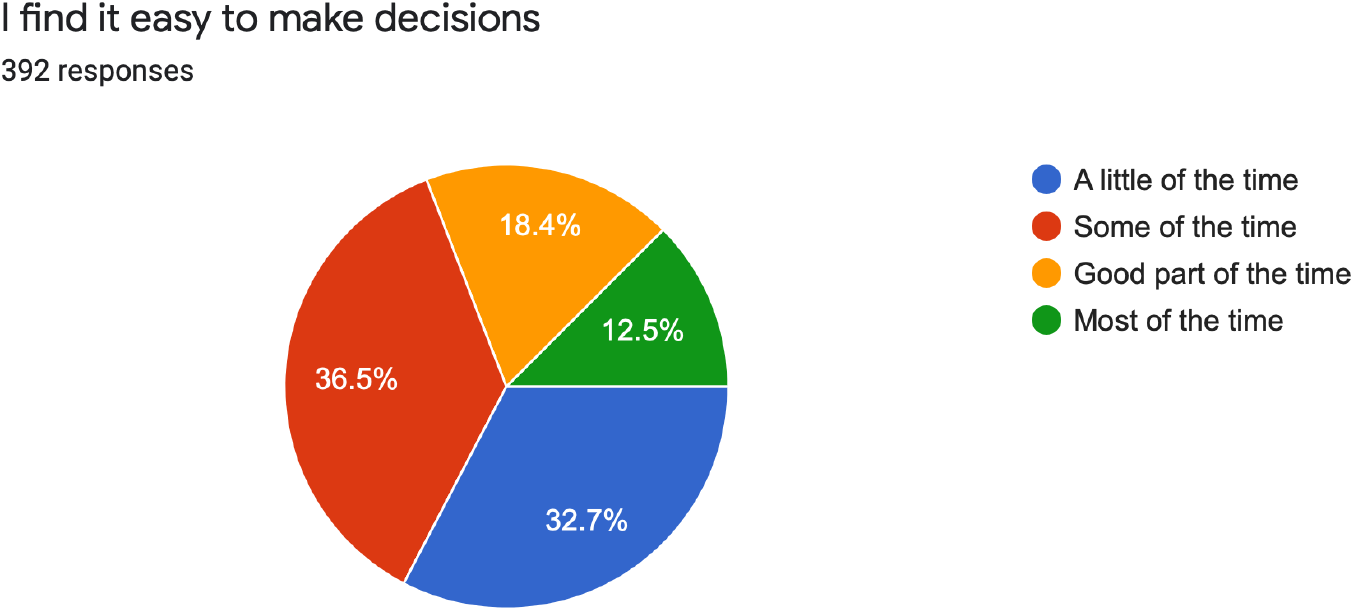

37. 27.3% of the respondents said that they felt they are useful and needed only for a little of the time while 24% of the respondents felt the same for good part of time.

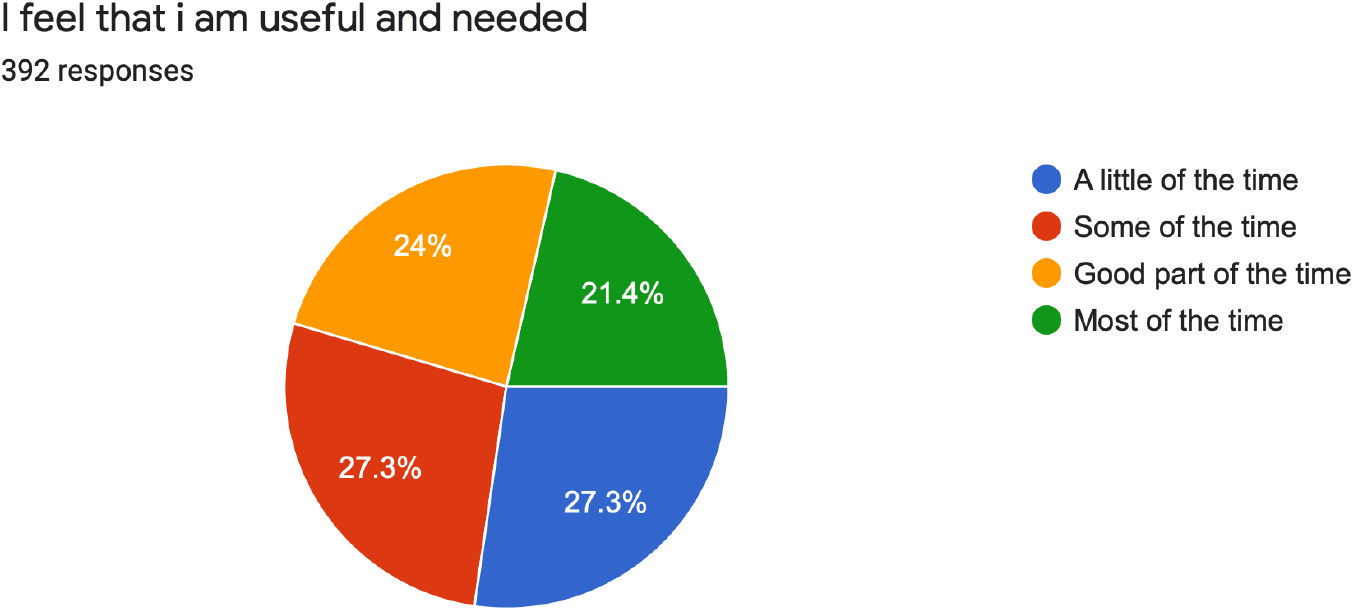

38. 30.1% of the participants felt their life is pretty full for a little of the time while 28.6% of the candidates felt the same for some of the time.

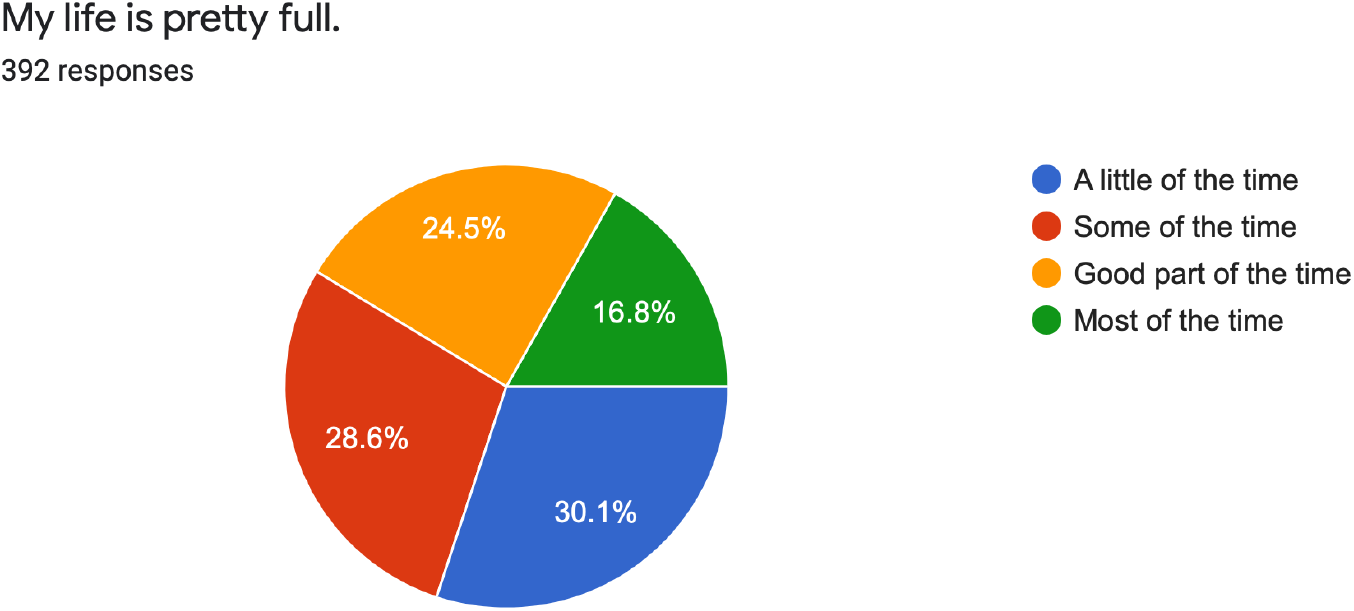

39. 12% of the participants reported that they fell others were better off if they were dead for most of the time. On the other hand 10.2% of the candidates felt the same for a good part of time.

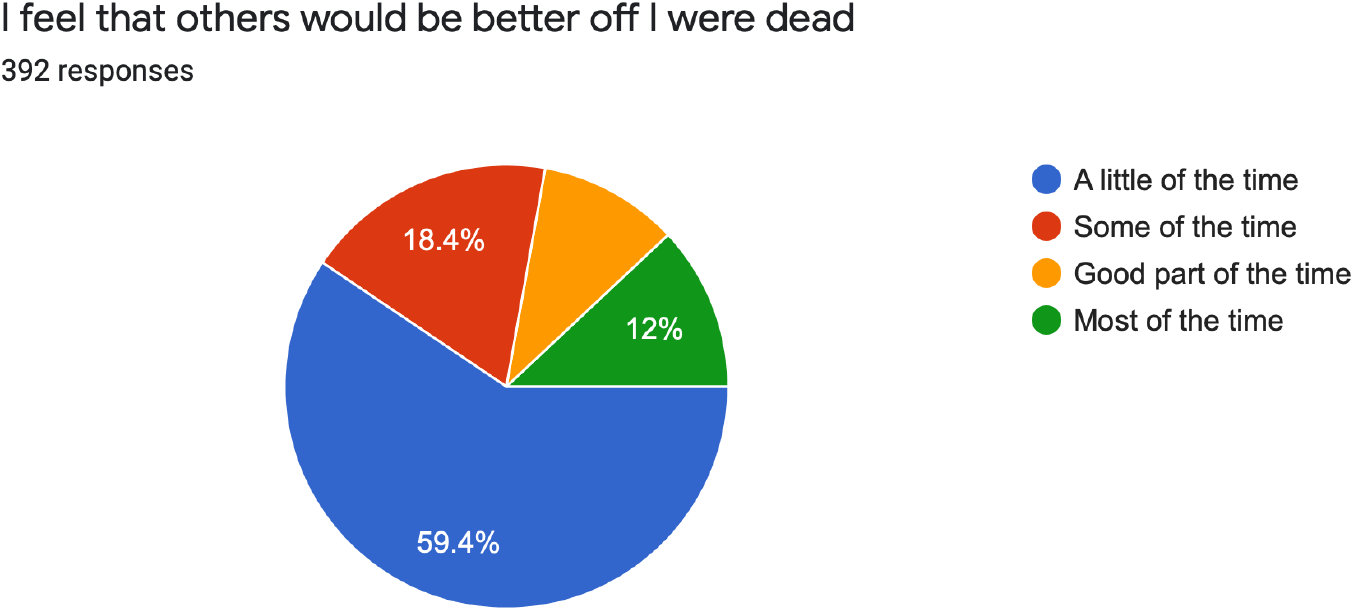

40. 18.4% of the candidates said they still enjoy the things they used to do for little of the time. While 28.1% of the candidates said that they felt the same for some of the time.

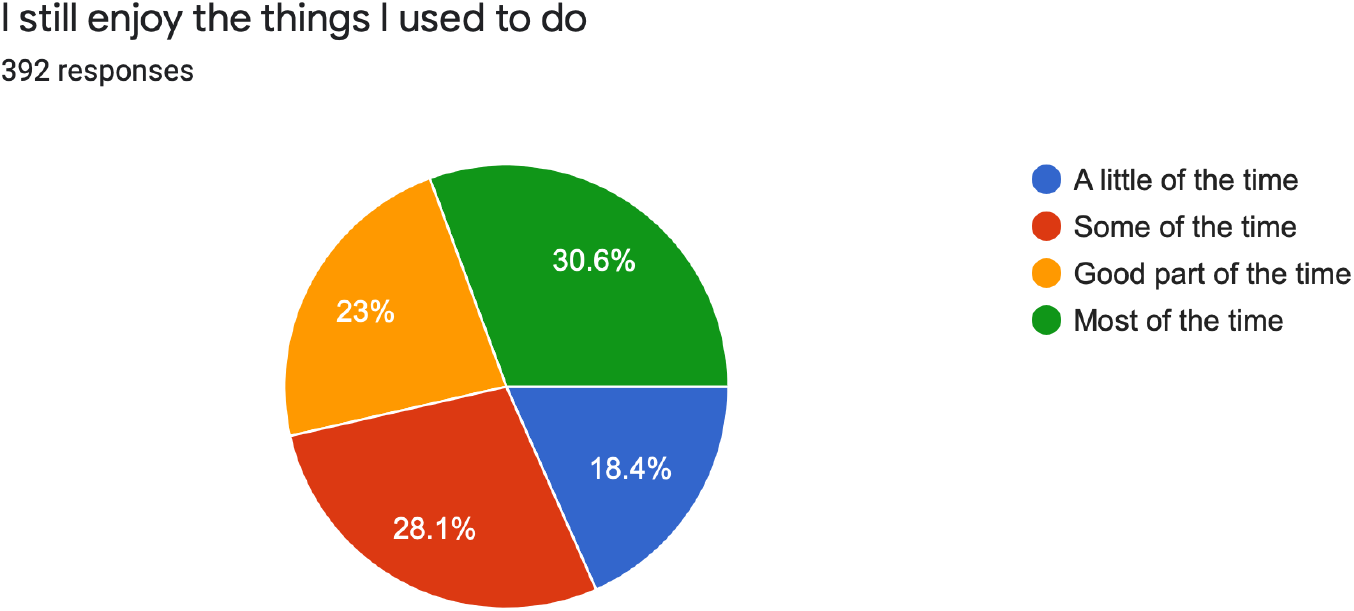

## DISCUSSION

We tried taking datas from students from all parts of the country in order to perform the study. A total of 392 students took part in our study. We have seen that 67 (17.1%) out of the total respondents were suffering from mild to moderate anxiety and 7 (1.79%) students had marked to severe anxiety. Coming to the depression part, 34 (8.6%) students had mild depression, while there was 1 (0.255%) responder having moderate and 1 (0.255%) having severe depression.

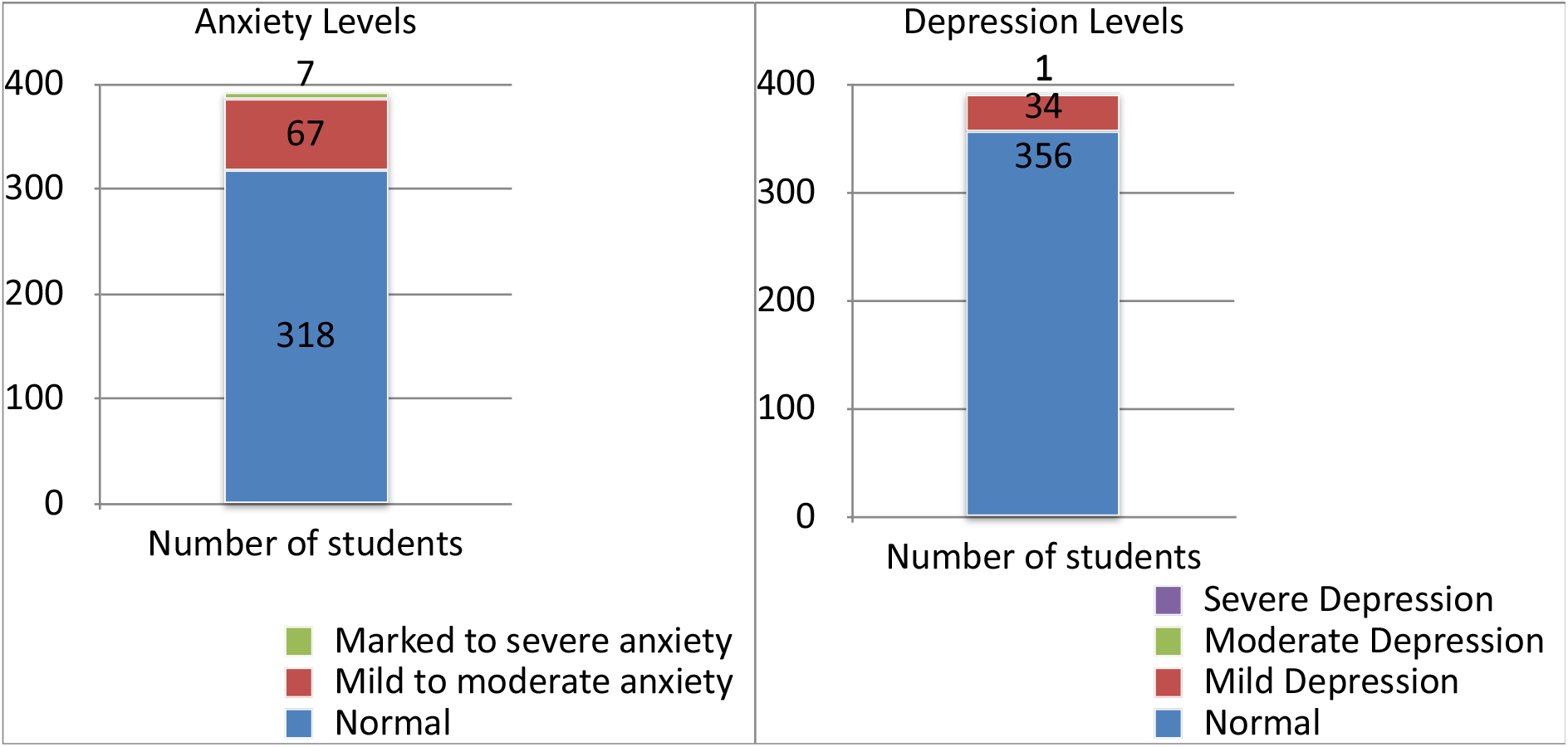

Our study had 168 (42.86%) female and 223 (56.89%) male respondents, while 2 candidates preferred not to disclose their gender. We also compared the relative anxiety and depression levels of male and female candidates and concluded that 22.03 % of females had significant levels of anxiety and 5.96% of the females had significant depression levels. On the other hand, 16.59% of the males had significant anxiety levels while 11.66% of the males had significant depression levels.

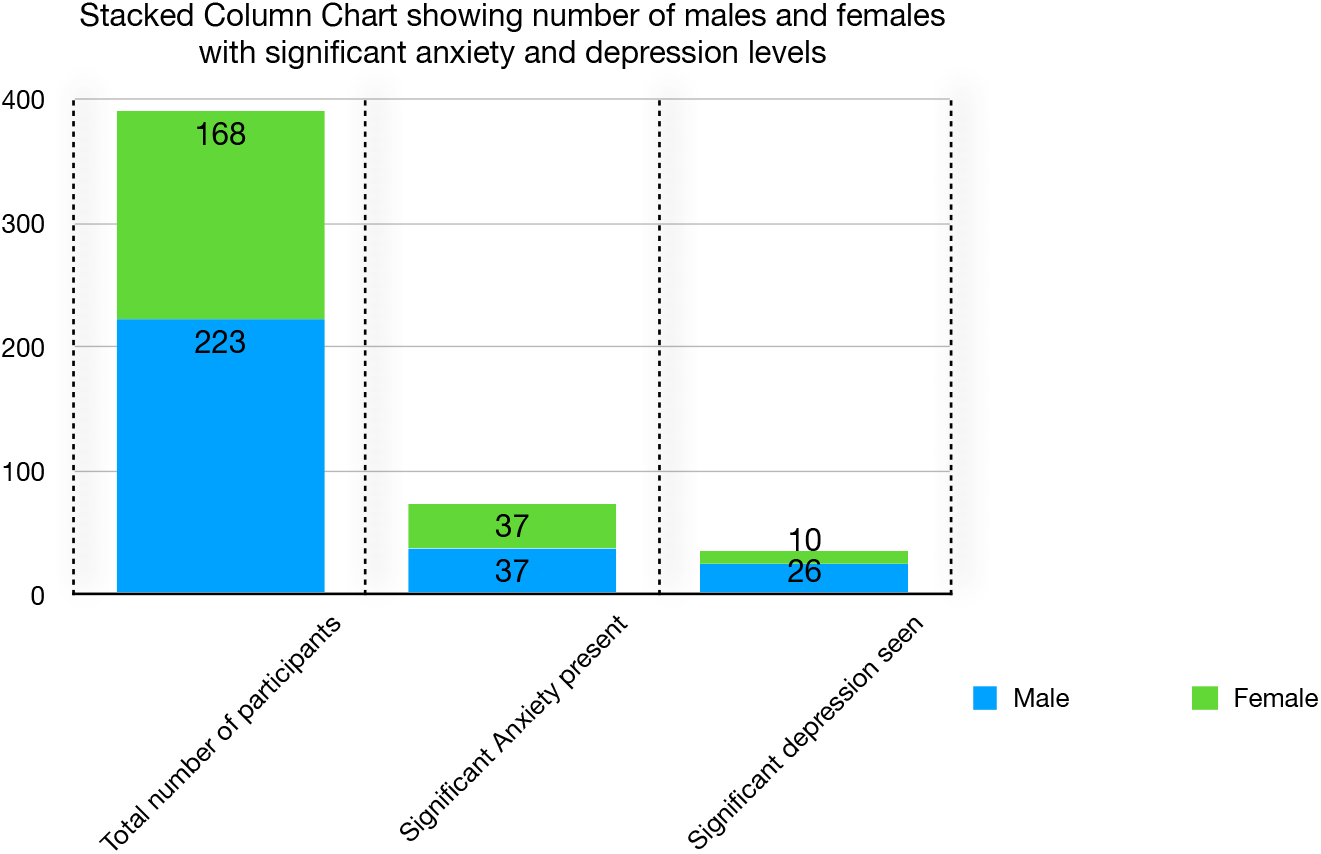

Out of the total candidates included in our study, 127 (32.4%) of them lived in rural areas whereas 265 (67.6%) were from urban areas. We compared the mental status in both livelihood areas and plotted the results in the following chart:

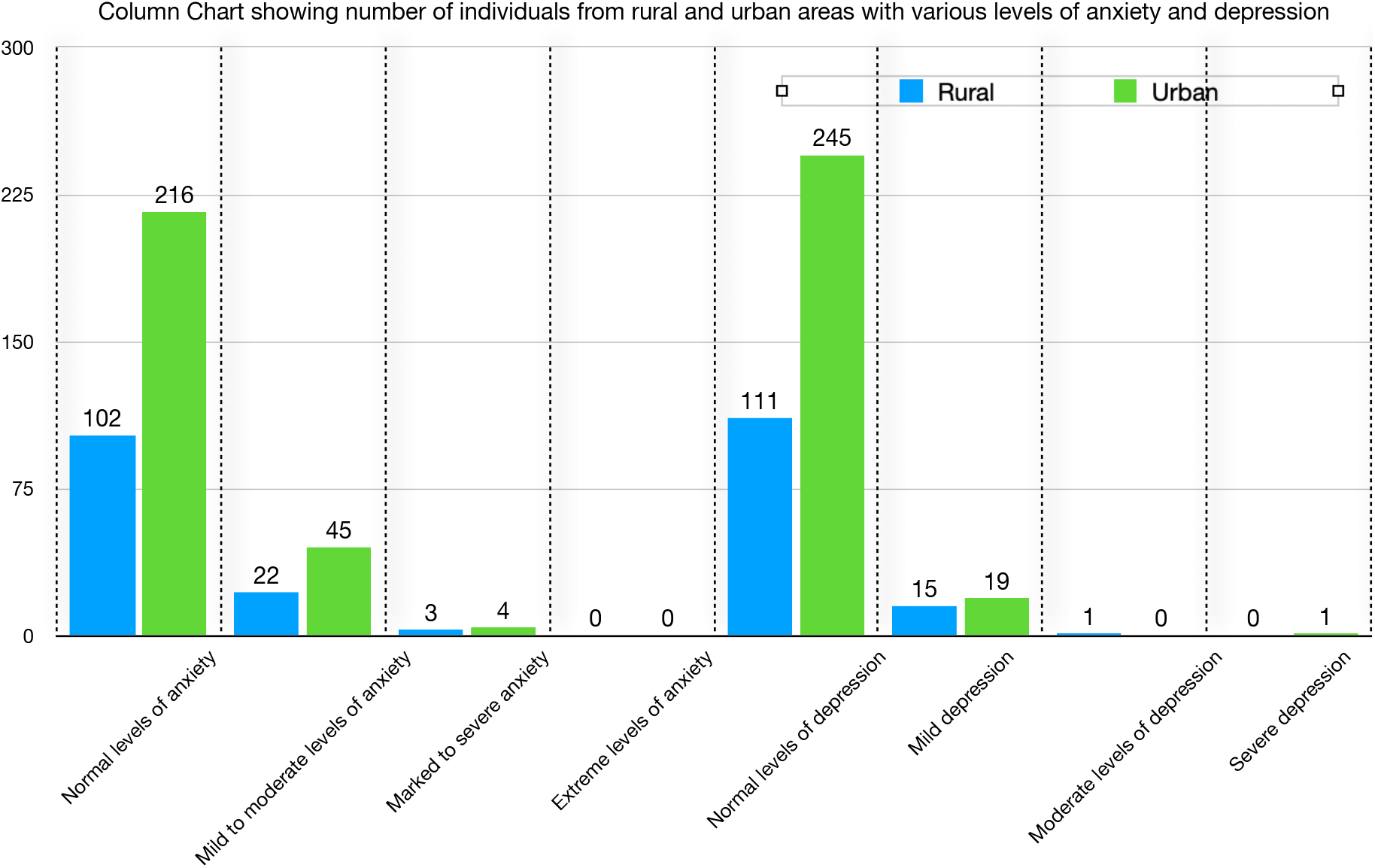

Our findings suggest a considerable negative impact of the COVID-19 pandemic on a variety of academic-, health-, and lifestyle-related outcomes. Our study will eventually help the students to be conscious about their psychological status and inculcate the rational perception towards Anxiety and Depression management.

While analysing the data, we also concluded that 2 of the respondents had marked to severe anxiety with mild depression and 8 candidates had mild to moderate anxiety with mild depression. Also, 1 candidate had marked to severe anxiety with moderate depression. On the other hand 1 student had marked to severe anxiety with severe depression which is a concern and required immediate attention. We mailed the respondent and requested him to visit psychologist, (as email was the sole way to communicate with the respondents).

We segregated the candidates according to their age group and sorted out the number of candidates having significant anxiety levels as per the scoring provided in the respective anxiety and depression scales, our conclusion is tabulated as following:

## CANDIDATES SORTED AS PER AGE GROUP AND EVALUATED AS PER ANXIETY SCALE

| **Age group** | **Total individuals** | **Normal Individuals** | **Mild to moderate anxiety** | **Marked to severe anxiety** | **Extreme anxiety** | **% of individuals having significant anxiety levels** |
| --- | --- | --- | --- | --- | --- | --- |
| 18-19 | 11 | 10 | 1 | 0 | 0 | 9.09% |
| 19-20 | 53 | 44 | 9 | 0 | 0 | 16.98% |
| 20-21 | 128 | 102 | 24 | 2 | 0 | 20.31% |
| 21-22 | 105 | 86 | 15 | 4 | 0 | 18.09% |
| 22-23 | 59 | 46 | 12 | 1 | 0 | 22.03% |
| 23-24 | 18 | 14 | 4 | 0 | 0 | 22.23% |
| 24-25 | 13 | 11 | 2 | 0 | 0 | 15.38% |
| 25-26 | 2 | 2 | 0 | 0 | 0 | —** |
| 26-27 | 2 | 2 | 0 | 0 | 0 | —** |
| 29 | 1 | 1 | 0 | 0 | 0 | —** |

REMARK:- We found that the candidates with age between 23 to 24 years were having highest prevalence of significant anxiety levels which is closely followed by candidates having age which lies between 22 years to 23 years.

## CANDIDATES SORTED AS PER AGE GROUP AND EVALUATED AS PER DEPRESSION SCALE

| Age group | Total individuals | Normal Individuals | Mildly depressed | Moderately depressed | Severely Depressed | % of individuals having significant anxiety levels |
| --- | --- | --- | --- | --- | --- | --- |
| 18-19 | 11 | 10 | 1 | 0 | 0 | 9.09% |
| 19-20 | 53 | 52 | 1 | 0 | 0 | 1.89% |
| 20-21 | 128 | 118 | 9 | 0 | 1 | 7.81% |
| 21-22 | 105 | 94 | 11 | 0 | 0 | 10.48% |
| 22-23 | 59 | 53 | 5 | 1 | 0 | 10.17% |
| 23-24 | 18 | 14 | 4 | 0 | 0 | 22.23% |
| 24-25 | 13 | 12 | 1 | 0 | 0 | 7.69% |
| 25-26 | 2 | 1 | 1 | 0 | 0 | 50% ** |
| 26-27 | 2 | 1 | 1 | 0 | 0 | 50% ** |
| 29 | 1 | 1 | 0 | 0 | 0 | — ** |

REMARK:- We found that candidates in the age group of 23-24 years had the maximum prevalence of depression. It was followed by candidates with ages between 21-22 years.
\*\* - age group don't have enough individuals to make a judgement

In light of the aforementioned projections of continued COVID-19 cases at the time of this writing and our findings, there is a need for immediate attention to and support for students and other vulnerable groups who have mental health issues.

Study shows that the pandemic has marked effect on the mental status of the students as the percentage of candidates with anxiety and depression cannot be ignored. Although there are some limitation in the study, we made it digital survey, so responders do have a smartphone and social media account. Due to the pandemic we were unable to reach the remote areas without electricity and internet. A proper awareness about the mental status and how to deal with it is highly needed among the students.

## Conclusion

Quite a significant number of candidates were suffering due to the pandemic situation. 17.091% were suffering from mild to moderate anxiety, 1.785% had marked to severe anxiety levels, (Constituting approximately 18.9% of the total). On the other hand, 8.673% of the students had mild depression, while 1 candidate (0.255%) had moderate depression and 1 (0.255%) had severe depression, (Constituting approximately 9.20% of the total)

## Data Availability

All data produced in the present study are available upon reasonable request to the authors

## Funding

NIL

## Conflict of Interest

No

## Consent of Respondents

Taken

## Contribution of Authors

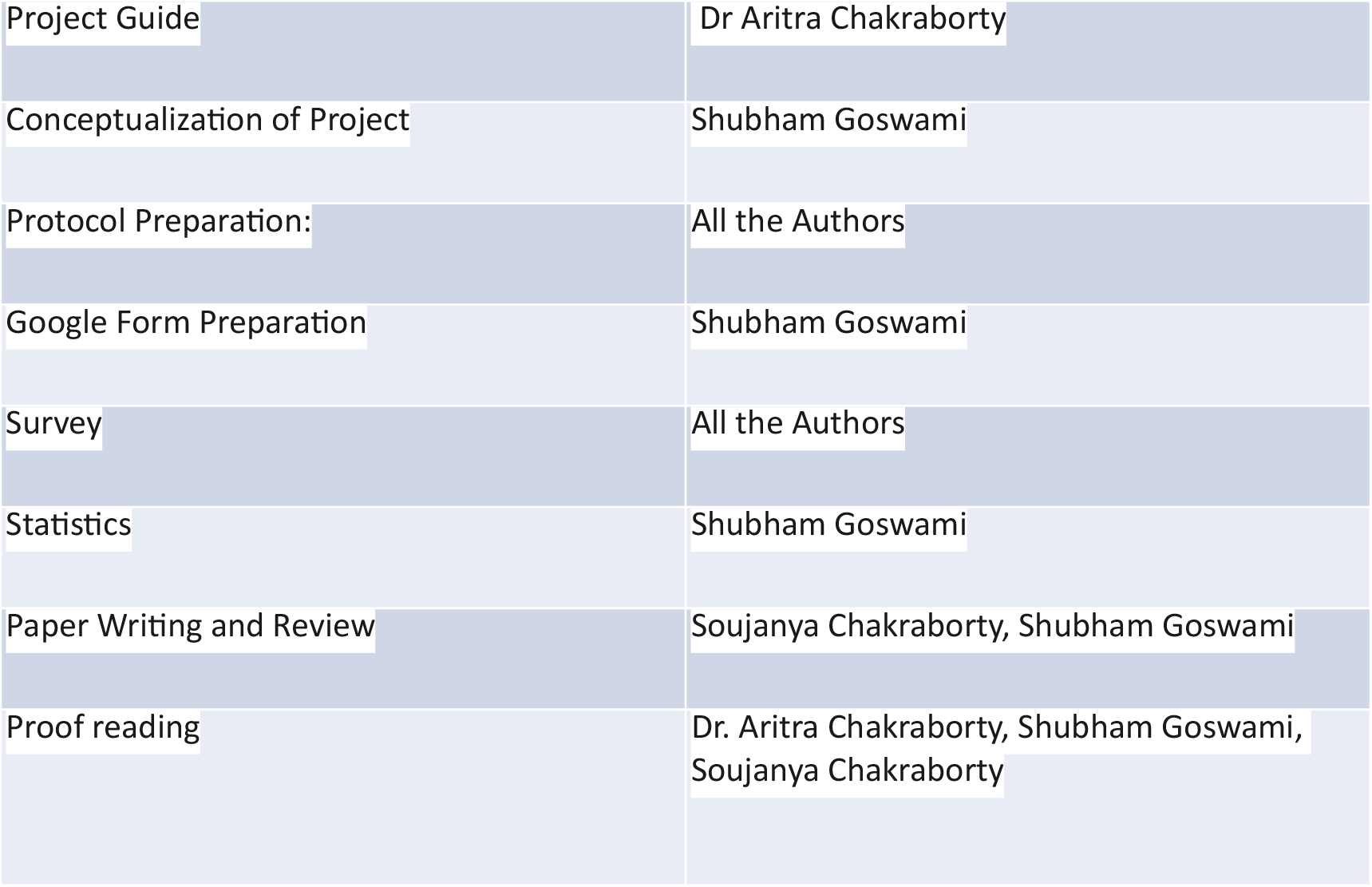

## Data Availability Statement

The data underlying this article will be shared on reasonable request to the corresponding author.

## Notes

### Competing Interest Statement

The authors have declared no competing interest.

### Funding Statement

This study did not receive any funding

### Author Declarations

Ethics committee of Bankura Sammilani Medical College gave ethical approval for this work

